# Meta-Analysis of Genome-Wide Association Studies of Hoarding Symptoms in 27 537 Individuals

**DOI:** 10.1101/2022.06.19.22276077

**Authors:** Nora I. Strom, Dirk J.A. Smit, Talisa Silzer, Conrad Iyegbe, Christie L. Burton, René Pool, Mathieu Lemire, James J. Crowley, Jouke-Jan Hottenga, Volen Z. Ivanov, Henrik Larsson, Paul Lichtenstein, Patrik Magnusson, Christian Rück, Russell Schachar, Hei Man Wu, Sandra M. Meier, Jennifer Crosbie, Paul D. Arnold, Manuel Mattheisen, Dorret I. Boomsma, David Mataix-Cols, Danielle Cath

## Abstract

Hoarding Disorder (HD) is a mental disorder characterized by persistent difficulties discarding or parting with possessions, often resulting in cluttered living spaces, distress, and impairment. Its etiology is largely unknown, but twin studies suggest that it is moderately heritable. In this study, we pooled phenotypic and genomic data from seven international cohorts (N = 27 537 individuals) and conducted a genome wide association study (GWAS) meta-analysis of parent- or self-reported hoarding symptoms (HS). We followed up the results with gene-based and gene-set analyses, as well as leave-one-out HS polygenic risk score (PRS) analyses. To examine a possible genetic association between hoarding symptoms and other phenotypes we conducted cross-trait PRS analyses. Though we did not report any genome-wide significant SNPs, we found a significant contribution of common genetic factors to HS, as indicated by substantial SNP-based twin-heritability estimates ranging between 26% and 48% and a SNP-heritability of 11% for one sub-cohort. Cross-trait PRS analyses showed that the genetic risk for schizophrenia and autism spectrum disorder were significantly associated with hoarding symptoms. We also found suggestive evidence for an association with educational attainment. There were no significant associations with other phenotypes previously linked to HD, such as obsessive-compulsive disorder, depression, anxiety, or attention-deficit hyperactivity disorder. To conclude, we found that HS are heritable, confirming and extending previous twin studies but we had limited power to detect any genome-wide significant loci. Much larger samples will be needed to further extend these findings and reach a “gene discovery zone”. To move the field forward, future research should not only include genetic analyses of quantitative hoarding traits in larger samples, but also in samples of individuals meeting strict diagnostic criteria for HD, and more ethnically diverse samples.

## 1. Introduction

Hoarding Disorder (HD) is one of the most recent mental disorders to be included in the DSM-5 (DSM-5, 2013) and ICD-11 (World Health Organization, 2019). Individuals with HD experience persistent difficulties parting with possessions, regardless of their value, due to a perceived need to save the items and distress associated with discarding them. This results in the accumulation of possessions that clutter active living areas and substantially compromise their intended use, causing clinically significant distress or impairment. Most people with HD also excessively acquire items that they do not need and experience distress if they are unable or are prevented from acquiring items (excessive acquisition specifier). Critically, these symptoms are not attributable to another medical or mental disorder, such as obsessive-compulsive disorder (OCD), psychosis or dementia (DSM-5; ICD-11).

The prevalence of HD in the population is estimated to be approximately 1% to 2.5% for both men and women (Ivanov et al., 2013, 2017a; Nordsletten et al., 2013; Postlethwaite et al., 2019), but a much larger proportion of the population experience symptoms at various levels of severity, with estimates up to 6.7% (Mathews et al., 2014) and 9% (Burton et al., 2018) in some studies. It is widely believed that the liability to hoarding symptoms (HS) is continuous in the general population, with clinically relevant HD at the extreme end of the spectrum (Timpano et al., 2013).

HS typically appear in early-to-mid-adolescence and, in contrast to many other psychiatric disorders, symptom severity increases with age (Ayers et al., 2010; Cath et al., 2017; Grisham et al., 2006; Ivanov et al., 2021). Psychiatric comorbidity is common in HD, with up to 70% of individuals having at least one additional disorder, most commonly anxiety and/or depression (DEP, Frost et al., 2011; Pertusa et al., 2008). Attentional problems are also common in individuals with HD (Frost et al., 2011; Fullana et al., 2013; Hartl et al., 2005; Tolin & Villavicencio, 2011).

The etiology of HD is largely unknown, though likely to be multifactorial in nature and related to a complex interplay of genetic, neurobiological, and psychosocial factors. Family studies have consistently shown that HS run in families (Frost & Gross, 1993; Pertusa et al., 2008; Samuels et al., 2002, 2007; Steketee et al., 2015). Population-based twin studies have estimated the heritability of hoarding symptoms based on self-report questionnaires (Iervolino et al., 2009, 2011; Ivanov et al., 2013, 2017a; López-Solà et al., 2014; C. A. Mathews et al., 2014; Taylor et al., 2010). In adults, heritability estimates range from 26% to 49%; the remaining variance was due to unique environmental factors and measurement error, whilst shared environmental factors appear to play a negligible role. In young people, large population-based samples of twins (N = 3 974 twins, Ivanov et al., 2013; N = 25 523, Ivanov et al., 2017), Ivanov et al. reported higher heritability of hoarding symptoms amongst 15-year-old boys than in girls (33% vs 17%) and significant shared environment influences (22%) among female twins only, while Burton et al. (2018) reported a heritability of 61% with no shared environment effect (221 twin pairs). Thus, it is possible that genetic and environmental influences on hoarding symptoms change across development, with shared environmental factors being more important in young people (particularly in girls).

Linkage and GWAS studies of HS have been rare thus far and have largely been conducted in small samples of OCD or Tourette syndrome patients. Candidate gene studies in individuals with OCD have suggested (largely non-replicated) associations between HS and a number of candidate variants (Alonso et al., 2008; Lochner et al., 2005; Sinopoli et al., 2020; Timpano et al., 2011; Wendland et al., 2009). Three previous modestly sized genome-wide linkage studies of HS in OCD or Tourette syndrome samples resulted in either no significant or conflicting results (Liang et al., 2008; Samuels et al., 2007; Zhang et al., 2002). One study in OCD patients found linkage between HS and a region on chromosome 14q23-32 (Samuels et al., 2007), and another linkage study in OCD patients found evidence for interaction with a region on chromosome 9q that houses *SLC1A1*, a glutamate transporter gene (Liang et al., 2008). One GWAS of OCD symptom dimensions reported *SETD3*, a gene highly expressed in the brain and involved in apoptotic processes and transcriptomic changes, to be associated with HS (Alemany-Navarro et al., 2020). Another GWAS focused on HS in a British twin cohort (Perroud et al., 2011). The sample included 3 304 twins from the TwinsUK cohort, predominantly female (91.8%), with a mean age of 56.8 years. All participants completed the Hoarding Rating Scale-Self-Report (HRS-SR; Tolin et al., 2010), a brief self-administered instrument consisting of five items (clutter, difficulty discarding, excessive acquisition, distress, and impairment). While no genome-wide significant loci were identified, two genomic loci on chromosomes 5 and 6 showed suggestive evidence for association with HS.

Larger samples are needed to increase power to detect significant genetic effects. Therefore, the current study aimed to conduct a GWAS meta-analysis of several large international cohorts from Sweden, the Netherlands, England, and Canada that included parent- or self-report hoarding scale data. We pooled data from seven population-based cohorts that together include 27 537 individuals (including 6 981 twin-pairs), representing a more than 8-fold increase in sample size compared to the previous study by Perroud et al (2011). We followed up the results with gene-based and gene-set analyses, as well as leave-one-out hoarding symptom polygenic risk score (PRS) analyses and cross-trait PRS analyses to examine a possible genetic association between other phenotypes and HS.

## 2. Materials and Methods

### 2.1. Cohorts & phenotype assessment

The Hoarding Symptom (HS) GWAS meta-analysis included individuals from seven different European-ancestry cohorts. Four cohorts are part of the Swedish Twin Registry (STR), namely different age groups of the *Child and Adolescent Twin Study in Sweden* (CATSS15, CATSS18, and CATSS24), and the *Young Adult Twins in Sweden Study* (YATSS). The other cohorts are from the Netherlands Twin Register (NTR), Spit for Science (SfS), and TwinsUK (see Supplementary Material for detailed descriptions). Data from the TwinsUK cohort were included in a previous GWAS (Perroud et al 2011). The cohorts are all population-based, predominantly including twins (except SfS), with a mean age-range between 11 and 44 (see Table 1). Participants, or their parents, answered one of two questionnaires assessing HS.

**Table 1:** Overview of cohorts included in the GWAS meta-analysis of HS. For each individual cohort included in the HS meta-analysis (STR-CATSS15, STR-CATSS18, STR-CATSS24, STR-YATSS, NTR, SfS, TwinsUK), the table lists the total sample size included (N), the number of monozygotic twin pairs (N MZ twin pairs), the number of dizygotic twin pairs (N DZ twin pairs), the number of siblings (N siblings), the number of parents (N parents), the percentage of females and males in the total N (% females (males)), and the mean age with standard deviations (SD). Twins where only one twin participated were not counted as twins. NTR twin pairs include 10 multiplets, remaining individuals also include 210 spouses. Note that CATSS samples were later pooled across the three cohorts (CATSS15, CATSS18, CATSS24) for GWAS analysis, depending on the platform they were genotyped on (GSA, PsychChip).

|  | STR-CATSS15 | STR-CATSS18 | STR-CATSS24 | STR YATSS | NTR | SfS | TwinsUK |
| --- | --- | --- | --- | --- | --- | --- | --- |
| **N** | 3 605 | 3 286 | 2 313 | 2 947 | 6 725 | 5 218 | 3 443 |
| **N MZ twin pairs** | 241 | 256 | 191 | 552 | 852 | - | 317 |
| **N DZ twin pairs** | 1 219 | 1 137 | 685 | 348 | 408 | - | 775 |
| **N siblings** | - | - | - | - | 644 | - | - |
| **N parents** | - | - | - | - | 2 062 | - | - |
| **% females** | 50% | 56% | 57% | 63% | 66% | 48% | 92% |
| **Mean age ± SD** | 15.47 ± 0.36 | 18.56 ± 0.33 | 23.84 ± 0.32 | 23.93 ± 1.78 | 41.4 ± 15.19 | 10.92 ± 2.79 | 56.7% ± 12.6 |

In STR, NTR, and TwinsUK, HS were assessed using four to five items of the Hoarding Rating Scale Self-Report (HRS-SR; Tolin et al., 2008), while in SfS parent- or self-reported hoarding traits were assessed using 2 items from the Toronto Obsessive Compulsive Scale (TOCS), a 21-item questionnaire described elsewhere (Burton et al., 2021; Park et al., 2016; see Supplementary Table S1 for questionnaire details). To summarize across HRS-SR items, four items of the HRS-SR were used to calculate a one-factor model using a latent variable analysis with the R package lavaan (Rosseel, 2012). In case an individual was missing one item, the mean of the remaining three items was used to impute the missing value. If more than one item was missing, the individual was removed from the analysis. The two SfS Hoarding items were summed and standardized into a Z-score. To ensure reliable and valid symptom reporting, SfS participants <12 years of age with self-reported HS were excluded. Distribution of the item and total raw hoarding scores are shown in Supplementary Figures S1-S6, distribution of the one-factor model scores (STR, NTR, and TwinsUK) and standardized scores (SfS) are shown in Supplementary Figures S7-S10.

### 2.2. Genome-wide association analysis

All participants were genotyped on SNP-arrays based on DNA from saliva or blood. One part of the STR-CATSS samples was genotyped on the PsychChip genotyping array (N = 8 598), another part was genotyped on the GSA genotyping array (N = 606). For the GWAS analyses STR-CATSS cohorts (CATSS15, CATSS18, CATSS24) were pooled over each genotyping platform (GSA, PsychChip), forming two separate CATSS datasets (STR-CATSS-GSA and STR-CTASS-PC). Each of the six datasets (STR-CATSS-GSA, STR-CATSS-PC, STR-YATSS, NTR, SfS, and TwinsUK) underwent stringent quality-control (QC), including the removal of non-European ancestry outliers based on PCA and imputation using the HRC (STR, NTR) or the 1000G (SfS, TwinsUK) reference sets (see Supplementary Material for more details). After genotyping, quality control, and imputation of each cohort, STR included 12 151, NTR 6 725, SfS 5 218, and TwinsUK 3 443 (total N = 27 537) individuals with complete genotypic and phenotypic information.

A linear mixed modeling GWAS was conducted within each cohort using GCTA-fastGWA (Yang et al., 2011). For STR, NTR, and TwinsUK a sparse Genetic Relatedness Matrix (GRM) was calculated and the first 10 principal components, sex, age, age squared, and genotyping batches were used as covariates. In a sparse GRM all off-diagonal values below 0.05 are set to For SfS, analyses were performed on unrelated individuals; the first enrolled sibling from each family was selected for further analysis. GCTA-fastGWA linear regression was performed with a full GRM and sex, age, respondent (parent vs. child reporting), genotyping array, principal components 1-3 and projected principal components 1-3 (see supplement) as covariates.

Next, the resulting GWAS summary statistics were cleaned and harmonized. All variants were filtered on minor allele frequency (MAF) > 1%, and imputation-quality score > 0.8. All datasets were aligned to the HRC-reference. In case alleles were reported on different strands, they were flipped to the orientation in the HRC reference. Strand ambiguous A/T and C/G SNPs were removed if their MAF was >= 0.4. Remaining ambiguous SNPs were strand aligned by comparing MAF to the HRC reference (McCarthy et al., 2016). We then used METAL (Willer et al., 2010) within the Rapid Imputation for COnsortias PIpeLIne (Ricopili) (Lam et al., 2020) to conduct an inverse variance weighted meta-analysis. The genomic control factor (Lambda and Lambda1000) was inspected for each individual cohort to detect any residual population stratification or systematic technical artifacts. Also, the LD score regression (LDSC) intercept was inspected as an alternative measure of test statistic inflation. The genome-wide significance threshold was set at 5×10^−8^.

### 2.3. Heritability

We used a restricted maximum likelihood (REML) approach in GCTA (S. H. Lee et al., 2011) to estimate the additive genetic variance (heritability) of each cohort separately, using the GRMs and covariates as used for the association analysis. We further used LDSC (Bulik-Sullivan et al., 2015) to calculate the SNP heritability of the HS GWAS meta-analysis. The SNP heritability in LDSC is based on the estimated slope from the regression of the SNP effect sizes from the GWAS on the LD score.

### 2.4. Gene-based and gene-set analyses

We carried out a Multi-marker Analysis of GenoMic Annotation (MAGMA) v1.08 (de Leeuw et al., 2015) as implemented in the web-based tool Functional Mapping and Annotation of Genome-Wide Association Studies (FUMA) v1.3.7 (Watanabe et al., 2017) to test genetic associations at the gene level for the combined effect of SNPs in or near protein coding genes. Gene-based p-values were computed by mapping SNPs to their corresponding gene(s) based on their position in the genome. Positional mapping was based on ANNOVAR annotations and the maximum distance between SNPs and genes was set to the default setting of 10 kb. Based on the results of gene analysis, competitive gene-set analysis was performed with default parameters. The 15 496 gene-sets were obtained from MsigDB v7.0, including ‘*Curated gene sets*’ consisting of nine data resources including KEGG, Reactome, and BioCarta, and *‘GO terms’* consisting of three categories (biological processes, cellular components, and molecular functions).

### 2.5. Cross-trait polygenic risk score (PRS) analyses

To explore the genetic relationship between HS and other phenotypes, we calculated a range of PRSs based on large-scale GWAS summary statistics. We selected mainly studies of psychiatric disorders, i.e. OCD (International Obsessive Compulsive Disorder Foundation Genetics Collaborative (IOCDF-GC) and OCD Collaborative Genetics Association Studies & OCGAS, 2017), DEP (Howard et al., 2019), schizophrenia (SCZ; Consortium et al., 2020), autism spectrum-disorder (ASD; Grove et al., 2019), attention-deficit-hyperactivity disorder (ADHD; Demontis et al., 2019), and educational attainment (EA; Lee et al., 2018). PRS were computed in PRSice2 for each cohort. The PRS scores were calculated as the weighted sum of the risk allele dosages at pre-selected p-value thresholds based on the reported thresholds in the primary publications (EA: P = 1; ADHD, ASD, OCD, SCZ: P = 0.01; DEP: P = 0.5). For STR, PRS analyses were conducted separately for the three datasets (STR-CATSS-GSA, STR-CATSS-PC, STR-YATSS) and were subsequently merged.

To evaluate the relationship between each PRS score and HS in every cohort, we employed generalized estimating equation (GEE) in R (STR, NTR, and TwinsUK). The GEE analysis accounts for the relatedness in the datasets. As SfS did not contain any related individuals, we carried out linear regression, as implemented within the PRSice2 pipeline. Again, the same covariates that were previously used in the respective GWASs were included.

PRS estimates per discovery phenotype were summarized across all target datasets by means of an inverse variance meta-analysis using the metagen package in R (R, 2017). We examined heterogeneity in PRS estimates across the cohorts with Cochran’s Q test (Cochran, 1950) and Higgin’s I^2^ (Higgins et al., 2003; Higgins & Thompson, 2002). Q is calculated as the weighted sum of the squared differences between individual cohort effects and the pooled effect across cohorts, with the weights being those used in the pooling method. The I^2^ statistic describes the percentage of variation across studies that is due to heterogeneity rather than sample variation and does, unlike Q, not inherently depend upon the number of measures included in the meta-analysis. Regardless of observed heterogeneity, we calculated a fixed effects model to evaluate the association of each PRS with HS. If there was considerable observed heterogeneity across study sites (I^2^ > 0.5 and/or P_Q_ < 0.05), we further calculated a random effects model.

### 2.6. Compatibility of cohorts

To identify if the summary statistics from any of the included cohorts substantially deviated from the others, we performed leave-one-out (LOO) GWAS meta-analyses and carried out a set of sensitivity analyses. With the replication module of the Ricopili pipeline, sign tests on the top SNPs (inclusion threshold of p = 0.0001, p = 0.00001, and p = 0.000001) were performed between each pair-wise combination of cohorts as well as between LOO meta-analyses and the left-out cohort to identify any cohort in which the GWAS results markedly deviated from the rest of the cohorts. Sign-tests allow for the quantification of independent genomic regions that have the same direction of effect in two separate summary statistics. The output, in form of a ratio, gives an estimate of the percentage of genomic regions with the same direction of effect in the two compared datasets. A ratio above 0.5 indicates convergence, while a ratio below 0.5 indicates divergence.

To evaluate the relationship between the PRS scores of each LOO GWAS and standardized HS scores in the left-out cohort, we conducted LOO PRS analyses, following the same procedure as for the cross-trait PRS analyses described above (see previous method-section on cross-trait PRS analyses for details).

## 3. Results

### 3.1. Genome-wide association results

The final dataset included 27 537 individuals with complete phenotypic and genotypic data and 6 541 342 autosomal SNPs. No significant inflation was observed (λ = 1.024, λ_1000_ = 1.001, LDSC intercept = 1.0173, see Supplementary Figure S11 for QQ plot). No SNP exceeded the genome-wide significance threshold (see Figure 1 for a Miami-plot including the Manhattan-plot of the GWAS in the upper panel). The SNPs with the lowest p-values (<1×10^−6^) were rs117321479 (P = 1.36×10^−7^) on chromosome 12, rs78426839 (P = 3.12×10^−7^) and rs7567224 (P = 7.70×10^−7^) on chromosome 2, and rs72927972 (P = 9.09×10^−7^) on chromosome 18 (see Supplementary Figures S12-S15 for regional association plots and forest plots). The region tagged by rs117321479 spans 57.6kb (LD r^2^ > 0.6) and entails the gene *SOX5*. The region tagged by rs78426839 spans 22.1kb (LD r^2^ > 0.6) and entails the genes *TUBA4B, DNAJB2, PTPRN, MIR153-1, RESP18*, and *DNPEP*. The region tagged by rs7567224 spans 24.40kb (LD r^2^ > 0.6) and entails the gene *CNTNAP5*, while the region tagged by rs78426839 spans 94.3kb (LD r^2^ > 0.6). Additionally, 19 independent SNPs with p < 1×10^−5^ were identified (see Supplementary Table S2 for a list of corresponding association results).

**Figure 1:**
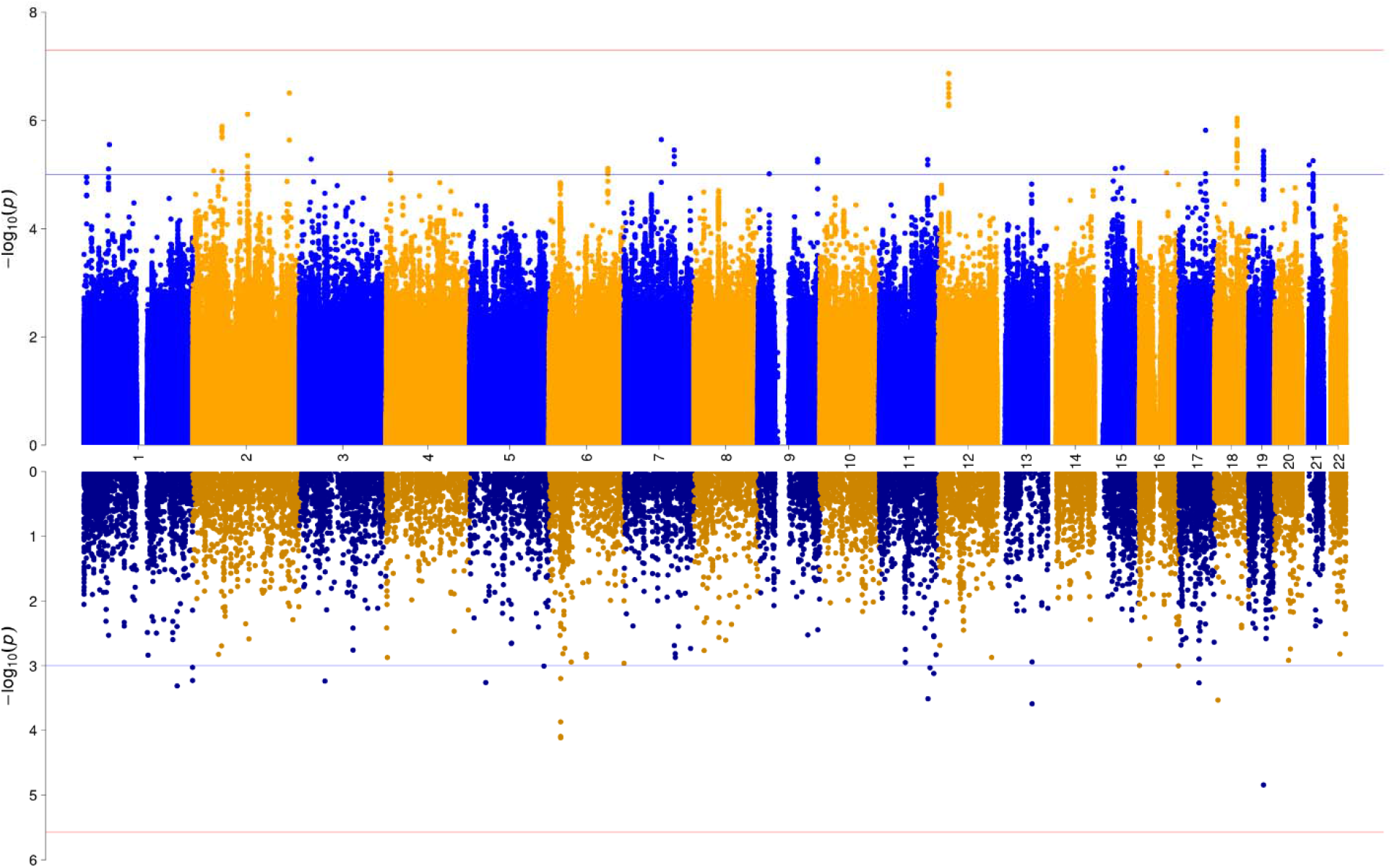
Miami plot of the association results from the GWAS meta-analysis (upper panel) and of the gene-wide association analysis (lower panel) of HS. The y-axes represent -log10 P values for the association of SNPs/genes with HS. The x-axis represents chromosomes 1 to 22. In the upper plot, the P-value threshold for genome-wide significance (5×10^−08^) is represented by the horizontal red line, suggestive significance (p = 1×10^−05^) by the blue line. In the lower panel, Bonferroni-corrected gene-wide significance (p = 2.682×10^−06^) is represented by the horizontal red line, suggestive gene-wide significance (p = 1×10^−03^) is indicated by the blue horizontal line.

### 3.2. Heritability

For the twin cohorts, the SNP-based additive genetic variance of HS, estimated based on the sparse genetic relatedness matrices, ranged between 0.26 (NTR) and 0.48 (TwinsUK), with estimates for the STR cohorts in between (STR-CATSS15: 0.47, STR-CATSS18: 0.29, STR-CATSS24: 0.35, STR-YATSS = 0.28). The SNP heritability for SfS (with only unrelated individuals) was 0.11 (SE = 0.057, P = 0.0303). Note that the SNP-heritability estimates based on the twin cohorts are largely driven by the twin resemblance (∼0.5 between DZ twins and siblings, 1.0 for MZ twins, and 0 between unrelated individuals), resulting in a SNP-heritability that mainly reflects the additive genetic variance from twin-modeling plus effects of shared environment, although the latter are known to play a negligible role in hoarding symptoms (Ivanov et al., 2017b; Zilhao et al., 2016). The SNP-based heritability estimate of the GWAS meta-analysis using LDSC resulted was 0.019 (SE = 0.016, Z = 1.18).

### 3.3. Gene-based analyses

We conducted gene-based tests to determine whether any protein-coding gene carries a load of common variation associated with HS. SNPs were mapped to 18 646 protein coding genes obtained from Ensembl build 85. No gene reached the Bonferroni-corrected significance threshold of p = 0.05/18 646 = 2.682×10^−6^ (see Figure 1 for a Miami-plot including the Manhattan-plot of the gene-based test in the lower panel, and Supplementary Figure S11 for a QQ-plot). Also, none of 15 483 tested gene-sets reached the Bonferroni-corrected significance threshold.

### 3.4. Cross-trait PRS analyses

To evaluate the genetic overlap between HS and other potentially related phenotypes of HS, we conducted PRS analyses. Publicly available summary statistics of OCD, DEP, SCZ, ASD, ADHD, and EA served as discovery datasets, with each HS cohort as the target dataset. GEE (STR, NTR, TwinsUK) and linear regression analysis (SfS) revealed Bonferroni-corrected significant (p < 0.05/7 = 0.00714) associations between HS and PRSs based on all discovery datasets, though not consistently across all target cohorts (see Supplementary Table S3). In a meta-analysis summarizing PRS results across all HS cohorts, the PRS for SCZ showed a significant association with HS (P_fixed_ = 2.43×10^−06^, P_random_ = 0.00422). When not taking the heterogeneity of the individual PRS estimates into account, the meta-analyzed PRSs of ASD and EA also showed significant associations with HS (ASD: P_fixed_ = 0.00426; EA: P_fixed_ = 1.15E^-05^). As the associations with the PRS of EA showed substantial (I^2^ = 0.76598, P_Q_ = 0.00504) heterogeneity, we further conducted random effects models to examine if the association with HS remains significant when accounting for this heterogeneity, resulting in a nominally significant association (EA: P_random_ = 0.00765). The associations with the PRS of DEP, OCD, and ADHD did not show any significant associations with HS in the meta-analysis (see Table 2 for all results).

**Table 2:** Depicted are results from the PRS meta-analysis for HS (leave-one-out), obsessive-compulsive disorder (OCD), depressive disorder (DEP), schizophrenia (SCZ), autism-spectrum disorder (ASD), attention-deficit hyperactivity disorder (ADHD), and educational attainment (EA) for pre-selected p-value thresholds (P_threshold_), meta-analyzed across all target datasets (STR, NTR, SfS, TwinsUK). As measures of heterogeneity of PRS associations across all target datasets, Higgin’s I^2^ statistic and the p-value for Cochran’s Q test (P_Q_) are reported. P_fixed_ and P_random_ list the p-values of a fixed and a random-effects model, respectively. A random effects model was only calculated if there was substantial (I^2^ > 0.5 and/or P_Q_ < 0.05) heterogeneity across the datasets. The effective sample size (N_effective_) is summed over the effective sample size of every target dataset. For STR, NTR, and TwinsUK the effective N was determined based on the actual N (including family members) weighted by the ratio of the squared SE from the GEE sandwich-corrected model and the naive model (no correction), for SfS the total sample N was used. Bonferroni-corrected significant p-values (< 0.05/7 = 0.00714) are in bold.

| Discovery | P<sub>threshold</sub> | N<sub>effective</sub> | Heterogeneity |  | P<sub>fixed</sub> | P<sub>random</sub> |
| --- | --- | --- | --- | --- | --- | --- |
|  |  |  | I<sup>2</sup> | P<sub>Q</sub> |  |  |
| Hoarding Symptoms (Leave-one-out) |  |  |  |  |  |  |
| HS | 0.5 | 25 388.59 | 0 | 0.9247 | 0.5525 |  |
| Cross-trait |  |  |  |  |  |  |
| OCD | 0.1 | 25 465.35 | 0 | 0.62865 | 0.67419 |  |
| DEP | 0.05 | 25 605.46 | 0.82711 | 0.00060 | 0.01499 | 0.58614 |
| SCZ | 0.1 | 25 547.3 | 0.44951 | 0.14168 | 2.43E<sup>-06</sup> |  |
| ASD | 0.1 | 25 976.75 | 0.26663 | 0.25184 | 0.00426 |  |
| ADHD | 0.1 | 25 937.77 | 0.55477 | 0.08073 | 0.21301 | 0.81557 |
| EA | 1 | 25 307.06 | 0.76598 | 0.00504 | 1.15E<sup>-05</sup> | 0.00765 |

### 3.5. Compatibility of cohorts

No genome-wide significant heterogeneity was observed in the HS GWAS meta-analysis (see Supplementary Figure S16 for Manhattan-plot and QQplot of the heterogeneity test). A range of sensitivity analyses, including LOO GWAS analyses, and subsequent sign-test analyses and LOO PRS analyses provided further evidence that there was no systematic and substantial heterogeneity across the different cohorts.

First, we conducted sign tests at three different p-value thresholds (1×10^−06^, 1×10^−05^, 1×10^−04^) between each pair-wise combination of STR, NTR, SfS, and TwinsUK (see Supplementary Table S4). Second, to determine if there was any pronounced age-related effect in the STR or NTR data, we conducted sign-test analyses between LOO GWAS analyses of age-separated STR sub-cohorts (Supplementary Table S5), age-separated NTR sub-cohorts (Supplementary Table S6) and the respective left-out cohort. The STR cohorts were divided into four age groups, pertaining to their division into separate phenotyping rounds (CATSS15 mean age = 15.46; CATSS18 mean age = 18.56; CATSS24 mean age = 23.84; YATSS mean age = 23.93), while the NTR data was separated into three age groups (group1 < 30 years; group2 30-45 years; group3 >45 years). The sign test results at the p-value threshold of p = 1×10^−06^ identified very few independent genomic regions (0-6) and are therefore difficult to interpret. While the ratios of the sign tests for the other two p-value thresholds (p = 1×10^−05^ and p = 1×10^−04^) varied between 0.2 and 0.8, there is no apparent pattern indicating a systematic deviation of one cohort from the rest.

The LOO PRS analyses did not show a significant association between any of the and the HS score of the left-out cohort, likely indicating a low power for this analysis (see Supplementary Table S3 and Table 2 for results).

## 4. Discussion

With 27 537 included individuals we conducted the largest GWAS study of HS in the population to date. Although we could not report any genome-wide significant SNPs, we found a significant contribution of common genetic factors to HS as indicated by a substantial genetic SNP heritability of 11% (P = 0.0303) in one of our cohorts (SfS) with unrelated samples. It suggests that, with sufficient power, specific genetic variants that are associated with HS will be eventually identified. SNP-based heritability of the meta-analysis as calculated with LDSC was low (h^2^ = 0.019, SE = 0.016, Z = 1.18) and non-significant. We therefore did not conduct genetic correlation analyses as it is suggested to have a heritability Z-score of above 1.5 (optimal > 4) in order for the analysis to be meaningful (Bulik-Sullivan et al., 2015; Zheng et al., 2017). We also found a significant genetic variance component in the twin family cohorts, ranging from 28% to 48%. These heritability estimates may be largely driven by the relatedness between the samples and are indeed more comparable to pedigree-based twin-heritability estimates. Ivanov et al (2017) reported twin-heritability estimates of 41%, 31%, and 29% for the Swedish CATSS15, CATSS18, and YATSS cohorts, respectively, while in this study we report heritability estimates of 47%, 29%, and 28% for the same cohorts, respectively.

The region tagged by the SNP with the lowest p-value in our analysis (rs117321479) entails the gene *SOX5*, which is a member of the *SOX* family of transcription factors involved in the regulation of embryonic development and in the determination of cell fate (Wunderle et al., 1996; provided by RefSeq, Jul 2008). *SOX5* has been identified as a gene with a high pleiotropic effect on a broad spectrum of psychiatric disorders and has been associated especially with ASD, BIP, MD, and SCZ, and to a lesser degree also with OCD, TS, ADHD, and AN (Lee et al., 2019). Overall, *SOX5* has been genome-wide significantly associated with 64 phenotypes, spanning a wide range of domains including psychiatric (e.g. neuroticism; Watanabe et al., 2019), skeletal (e.g. height; Wood et al., 2014), reproductive (e.g. age at menarche; Day et al., 2017), metabolic (e.g. hip circumference, BMI; Watanabe et al., 2019), and environmental traits (e.g. household income (Watanabe et al., 2019), educational attainment (Lee et al., 2018)). Given this possible pleiotropy, it suggests that *SOX5* might be involved in susceptibility to general psychopathology.

By current standards (Howard et al., 2019; Consortium et al., 2020; Grove et al., 2019; Demontis et al., 2019), the size of our GWAS was modest and no significant effects were observed in the leave-one-sample-out analysis of PRS associated with HS, indicating that the lack of finding genome-wide hits is most likely attributable to a lack of power. We further observed a rather high number of individuals (28.8%) with zero scores on the hoarding rating scales, leading to a relatively low variance of symptom scores in our dataset, and our datasets included a high number of twins, both of which likely reduced the effective sample size and contributed to the power issue.

Another reason why we did not find a significant signal may lie in differences between the included study cohorts, beyond any heterogeneity that we were able to detect. Possible sources of heterogeneity across the datasets include age, instruments used to assess HS, or ascertainment of data. The SfS cohort stands out compared to the other cohorts with regards to age and questionnaire used. We addressed this issue by applying a framework of sensitivity and heterogeneity analyses. Neither in the genome-wide heterogeneity test, nor in the sign-tests or LOO PRS analyses did we observe a pattern that indicated a systematic deviation of one cohort from the rest. We therefore concluded that a meta-analysis of the cohorts is warranted and that the lack of signal is indeed most likely attributable to a lack of power. As most of the individuals included in the cohorts in this study were relatively young compared to the age when the full disorder debuts, there is the possibility that HS reported by younger individuals are the outcome of a somewhat different phenotype than HS reported at an older age.

Cross-trait PRS analyses showed significant results. The genetic risk for schizophrenia was significantly associated with HS, while we found suggestive evidence for an association with autism spectrum disorder and educational attainment. This suggests that a well-powered GWAS such as those used in the cross-trait PRS analyses (SCZ: N_effective_ = 214 576; ASD: N_effective_ = 44 367; EA: N_effective_ = 245 621) can pick up genetic signals that may be associated with hoarding symptoms. We consider this a further indicator that additional power is needed to obtain reliable signal in a quantitative trait GWAS for HS. The lack of significant associations with the most described comorbidities in HD (OCD, depression, anxiety, ADHD) was unexpected and possibly attributable to the currently modest size of the discovery samples.

Previous twin studies of HS have shown a clear role of genetic factors (Iervolino et al., 2009, 2011; Ivanov et al., 2013, 2017a; López-Solà et al., 2014; C. A. Mathews et al., 2014; Taylor et al., 2010). The current study confirmed these observations with a significant SNP-based heritability of HS. It is not entirely surprising that we did not discover any genome-wide significant associations, as other studies with similar sample sizes that investigated quantitative measures of symptoms also lacked power to discover associations (Ebejer et al., 2013; Middeldorp et al., 2016). Nevertheless, the quantitative ADHD symptom GWAS by Middeldorp et al. (2016) showed a high genetic correlation and strong concordance at individual loci with clinical ADHD (Demontis et al., 2019), thereby supporting the hypothesis that clinically diagnosed cases are the extreme of a quantitative symptom trait and further demonstrating the usefulness and importance of quantitative assessment of symptoms in the population.

A further limitation is that hoarding symptom scales may reflect heterogeneous disorders. It is known that many different psychiatric disorders can cause hoarding-like symptoms, such as schizophrenia, OCD, or severe depression. HD is essentially a diagnosis of exclusion (DSM-5, ICD-11). As in any study based on self-administered instruments, it was not possible to rule out these other causes of HS. It will further be of interest to determine the extent to which HS in the population and clinical HD share the same genetic susceptibility. However, no case-control GWAS of HD exists, nor is HD assessed in large datasets like UK BioBank (Bycroft et al., 2018; Sudlow et al., 2015), which is unfortunate given the relatively high prevalence of around 2.5% for HD (Postlethwaite et al., 2019) and high individual and societal cost (Frost, Steketee, & Williams, 2000a; Frost, Steketee, Williams, et al., 2000b; Carol A. Mathews et al., 2016). Thus, for the time being, the study of HS in the population may be the only feasible approach to understanding the genetics of HD.

To conclude, we found that HS are heritable, confirming and extending previous twin studies. Nevertheless, we had limited power to detect any genome-wide significant loci. Much larger samples will be needed to further extend our findings and reach a “gene discovery zone”. Further, additional samples should be more ethnically diverse to ensure that results are relevant to individuals of non-European ancestry (Fernández de la Cruz et al., 2016). Future research should include the collection of DNA samples from individuals with HS, as well as samples from strictly diagnosed HD patients.

## Data Availability

The meta-analyzed summary statistics will be made available via the Psychiatric Genomics Consortium Download page upon publication in a journal (https://www.med.unc.edu/pgc/download-results/).

## 5. Acknowledgements

We acknowledge The Swedish Twin Registry (STR) for data access.

The STR is managed by Karolinska Institutet and receives funding through the Swedish Research Council under the grant no 2017-00641. The computations/data handling were enabled by resources provided by the Swedish National Infrastructure for Computing (SNIC) at Uppmax partially funded by the Swedish Research Council through grant agreement no. 2018-05973.

The Netherlands Twin Register (NTR) warmly thanks all twin families for their participation.

NTR is supported by multiple grants from the Netherlands Organizations for Scientific Research (NWO) and Medical Research (ZonMW): Netherlands Twin Registry Repository (NWO 480-15-001/674); the Biobank-based integrative omics study (BIOS) funded by BBMRI-NL (NWO projects 184.021.007 and 184.033.111); the European Science Council (ERC) Genetics of Mental Illness (ERC Advanced, 230374, PI Boomsma); the Royal Netherlands Academy of Science Professor Award (PAH/6635) to D.I.B.; Rutgers University Cell and DNA Repository (NIMH U24 MH068457-06), the Avera Institute, Sioux Falls, South Dakota (USA) and the National Institutes of Health (NIH R01 HD042157-01A1). Part of the genotyping was funded by the Genetic Association Information Network (GAIN) of the Foundation for the National Institutes of Health and Grand Opportunity grants 1RC2 MH089951).

TwinsUK is funded by the Wellcome Trust, Medical Research Council, Versus Arthritis, European Union Horizon 2020, Chronic Disease Research Foundation (CDRF), Zoe Ltd, the National Institute for Health and Care Research (NIHR) Clinical Research Network (CRN) and Biomedical Research Centre based at Guy’s and St Thomas’ NHS Foundation Trust in partnership with King’s College London.

Spit for Science was supported by the Canadian Institutes of Health Research (R.J.S., MOP-93696 and P.D.A., MOP-106573). Dr. Arnold is supported by the Alberta Innovates Translational Health Chair in Child and Youth Mental Health.

## 7. Data availability

The meta-analyzed summary statistics will be made available via the Psychiatric Genomics Consortium Download page (https://www.med.unc.edu/pgc/download-results/).

Supplementary information is available at MP’s website.

## References

Alemany-Navarro, M., Cruz, R., Real, E., Segalàs, C., Bertolín, S., Rabionet, R., Carracedo, Á., Menchón, J. M., & Alonso, P. (2020). Looking into the genetic bases of OCD dimensions: a pilot genome-wide association study. Translational Psychiatry, 10(1), 1–16. https://doi.org/10.1038/s41398-020-0804-z

Alonso, P., Gratacòs, M., Menchón, J. M., Segalàs, C., González, J. R., Labad, J., Bayés, M., Real, E., De Cid, R., Pertusa, A., Escaramís, G., Vallejo, J., & Estivill, X. (2008). Genetic susceptibility to obsessive-compulsive hoarding: the contribution of neurotrophic tyrosine kinase receptor type 3 gene1. Genes, Brain and Behavior, 7(7), 778–785. https://doi.org/10.1111/J.1601-183X.2008.00418.X

Ayers, C. R., Saxena, S., Golshan, S., & Wetherell, J. L. (2010). Age at onset and clinical features of late life compulsive hoarding. International Journal of Geriatric Psychiatry, 25(2), 142–149. https://doi.org/10.1002/GPS.2310

Bulik-Sullivan, B. K., Loh, P.-R., Finucane, H. K., Ripke, S., Yang, J., Patterson, N., Daly, M. J., Price, A. L., & Neale, B. M. (2015). LD Score regression distinguishes confounding from polygenicity in genome-wide association studies. Nat Genet, 47(3), 291–295. https://doi.org/10.1038/ng.3211

Burton, C. L., Lemire, M., Xiao, B., Corfield, E. C., Erdman, L., Bralten, J., Poelmans, G., Yu, D., Shaheen, S. M., Goodale, T., Sinopoli, V. M., Askland, K. D., Barlassina, C., Bienvenu, O. J., Black, D., Bloch, M., Brentani, H., Camarena, B., Cappi, C., … Arnold, P. D. (2021). Genome-wide association study of pediatric obsessive-compulsive traits: shared genetic risk between traits and disorder. Translational Psychiatry 2021 11:1, 11(1), 1–10. https://doi.org/10.1038/s41398-020-01121-9

Burton, C. L., Park, L. S., Corfield, E. C., Forget-Dubois, N., Dupuis, A., Sinopoli, V. M., Shan, J., Goodale, T., Shaheen, S. M., Crosbie, J., Schachar, R. J., & Arnold, P. D. (2018). Heritability of obsessive–compulsive trait dimensions in youth from the general population. Translational Psychiatry, 8(1), 191. https://doi.org/10.1038/s41398-018-0249-9

Bycroft, C., Freeman, C., Petkova, D., Band, G., Elliott, L. T., Sharp, K., Motyer, A., Vukcevic, D., Delaneau, O., O’Connell, J., Cortes, A., Welsh, S., Young, A., Effingham, M., McVean, G., Leslie, S., Allen, N., Donnelly, P., & Marchini, J. (2018). The UK Biobank resource with deep phenotyping and genomic data. Nature 2018 562:7726, 562(7726), 203–209. https://doi.org/10.1038/s41586-018-0579-z

Cath, D. C., Nizar, K., Boomsma, D., & Mathews, C. A. (2017). Age-Specific Prevalence of Hoarding and Obsessive Compulsive Disorder: A Population-Based Study. The American Journal of Geriatric Psychiatry_J_: Official Journal of the American Association for Geriatric Psychiatry, 25(3), 245–255. https://doi.org/10.1016/J.JAGP.2016.11.006

Cochran, W. G. (1950). The comparison of percentages in matched samples. Biometrika, 37(3– 4), 256–266. https://doi.org/10.1093/BIOMET/37.3-4.256

Consortium, T. S. W. G. of the P. G., Ripke, S., Walters, J. T., & O’Donovan, M. C. (2020). Mapping genomic loci prioritises genes and implicates synaptic biology in schizophrenia. MedRxiv, 2020.09.12.20192922. https://doi.org/10.1101/2020.09.12.20192922

Day, F. R., Thompson, D. J., Helgason, H., Chasman, D. I., Finucane, H., Sulem, P., Ruth, K. S., Whalen, S., Sarkar, A. K., Albrecht, E., Altmaier, E., Amini, M., Barbieri, C. M., Boutin, T., Campbell, A., Demerath, E., Giri, A., He, C., Hottenga, J. J., … Perry, J. R. B. (2017). Genomic analyses identify hundreds of variants associated with age at menarche and support a role for puberty timing in cancer risk. Nature Genetics, 49(6), 834–841. https://doi.org/10.1038/NG.3841

de Leeuw, C. A., Mooij, J. M., Heskes, T., & Posthuma, D. (2015). MAGMA: Generalized Gene-Set Analysis of GWAS Data. PLoS Comput Biol, 11(4), 1–19. https://doi.org/10.1371/journal.pcbi.1004219

Demontis, D., Walters, R. K., Martin, J., Mattheisen, M., Als, T. D., Agerbo, E., Børglum, A. D., & Neale, B. M. (2019). Discovery of the first genome-wide significant risk loci for attention deficit/hyperactivity disorder. Nat Genet, 51(1), 63–75. https://doi.org/10.1038/s41588-018-0269-7

DSM-5. (2013). Diagnostic and statistical manual of mental disorders: DSM-5. 5 ed., 237–242. ISBN 978-0-89042-555–558. https://doi.org/https://doi.org/10.1176/appi.books.9780890425596

Ebejer, J. L., Duffy, D. L., Van Der Werf, J., Wright, M. J., Montgomery, G., Gillespie, N. A., Hickie, I. B., Martin, N. G., & Medland, S. E. (2013). Genome-wide association study of inattention and hyperactivity-impulsivity measured as quantitative traits. Twin Research and Human Genetics_J_: The Official Journal of the International Society for Twin Studies, 16(2), 560–574. https://doi.org/10.1017/THG.2013.12

Fernández de la Cruz, L., Nordsletten, A., & Mataix-Cols, D. (2016). Ethnocultural Aspects of Hoarding Disorder. Current Psychiatry Reviews. https://www.ingentaconnect.com/content/ben/cpsr/2016/00000012/00000002/art00005

Frost, R. O., & Gross, R. C. (1993). The hoarding of possessions. Behaviour Research and Therapy, 31(4), 367–381. https://doi.org/10.1016/0005-7967(93)90094-B

Frost, R. O., Steketee, G., & Tolin, D. F. (2011). Comorbidity in hoarding disorder. Depression and Anxiety, 28(10), 876–884. https://doi.org/10.1002/DA.20861

Frost, R. O., Steketee, G., & Williams, L. (2000). Hoarding: A community health problem. Health and Social Care in the Community, 8(4), 229–234. https://doi.org/10.1046/J.1365-2524.2000.00245.X

Frost, R. O., Steketee, G., Williams, L. F., & Warren, R. (2000). Mood, personality disorder symptoms and disability in obsessive compulsive hoarders: a comparison with clinical and nonclinical controls. Behaviour Research and Therapy, 38(11), 1071–1081. https://doi.org/10.1016/S0005-7967(99)00137-0

Fullana, M. A., Vilagut, G., Mataix-Cols, D., Adroher, N. D., Bruffaerts, R., Bunting, B., De Almeida, J. M. C., Florescu, S., De Girolamo, G., De Graaf, R., Haro, J. M., Kovess, V., & Alonso, J. (2013). Is ADHD in childhood associated with lifetime hoarding symptoms? An epidemiological study. Depression and Anxiety, 30(8), 741–748. https://doi.org/10.1002/DA.22123

Grisham, J. R., Frost, R. O., Steketee, G., Kim, H. J., & Hood, S. (2006). Age of onset of compulsive hoarding. Journal of Anxiety Disorders, 20(5), 675–686. https://doi.org/10.1016/J.JANXDIS.2005.07.004

Grove, J., Ripke, S., Als, T. D., Mattheisen, M., Børglum, A. D., & et al. (2019). Identification of common genetic risk variants for autism spectrum disorder. 51(3), 431–444. https://doi.org/10.1038/s41588-019-0344-8

Hartl, T. L., Duffany, S. R., Allen, G. J., Steketee, G., & Frost, R. O. (2005). Relationships among compulsive hoarding, trauma, and attention-deficit/hyperactivity disorder. Behaviour Research and Therapy, 43(2), 269–276. https://doi.org/10.1016/J.BRAT.2004.02.002

Higgins, J. P. T., & Thompson, S. G. (2002). Quantifying heterogeneity in a meta-analysis. Statistics in Medicine, 21(11), 1539–1558. https://doi.org/10.1002/SIM.1186

Higgins, J. P. T., Thompson, S. G., Deeks, J. J., & Altman, D. G. (2003). Measuring inconsistency in meta-analyses. BMJ, 327(7414), 557–560. https://doi.org/10.1136/BMJ.327.7414.557

Howard, D. M., Adams, M. J., Clarke, T. K., Hafferty, J. D., Gibson, J., Shirali, M., Coleman, J. R. I., Hagenaars, S. P., Ward, J., Wigmore, E. M., Alloza, C., Shen, X., Barbu, M. C., Xu, E. Y., Whalley, H. C., Marioni, R. E., Porteous, D. J., Davies, G., Deary, I. J., … McIntosh, A. M. (2019). Genome-wide meta-analysis of depression identifies 102 independent variants and highlights the importance of the prefrontal brain regions. Nature Neuroscience 2019 22:3, 22(3), 343–352. https://doi.org/10.1038/s41593-018-0326-7

Iervolino, A. C., Perroud, N., Fullana, M. A., Guipponi, M., Cherkas, L., Collier, D. A., & Mataix-Cols, D. (2009). Prevalence and heritability of compulsive hoarding: a twin study. The American Journal of Psychiatry, 166(10), 1156–1161. https://doi.org/10.1176/APPI.AJP.2009.08121789

Iervolino, A. C., Rijsdijk, F. V., Cherkas, L., Fullana, M. A., & Mataix-Cols, D. (2011). A multivariate twin study of obsessive-compulsive symptom dimensions. Archives of General Psychiatry, 68(6), 637–644. https://doi.org/10.1001/ARCHGENPSYCHIATRY.2011.54

International Obsessive Compulsive Disorder Foundation Genetics Collaborative (IOCDF-GC) and OCD Collaborative Genetics Association Studies, & OCGAS. (2017). Revealing the complex genetic architecture of obsessive-compulsive disorder using meta-analysis. Mol Psychiatry, 23(5), 1181–1188. https://doi.org/10.1038/mp.2017.154

Ivanov, V. Z., Mataix-Cols, D., Serlachius, E., Brander, G., Elmquist, A., Enander, J., & Rück, C. (2021). The developmental origins of hoarding disorder in adolescence: a longitudinal clinical interview study following an epidemiological survey. European Child & Adolescent Psychiatry, 30(3), 415–425. https://doi.org/10.1007/S00787-020-01527-2

Ivanov, V. Z., Mataix-Cols, D., Serlachius, E., Lichtenstein, P., Anckarsäter, H., Chang, Z., Gumpert, C. H., Lundström, S., Långström, N., & Rück, C. (2013). Prevalence, Comorbidity and Heritability of Hoarding Symptoms in Adolescence: A Population Based Twin Study in 15-Year Olds. PLoS ONE, 8(7), 1–7. https://doi.org/10.1371/journal.pone.0069140

Ivanov, V. Z., Nordsletten, A., Mataix-Cols, D., Serlachius, E., Lichtenstein, P., Lundström, S., Magnusson, P. K. E., Kuja-Halkola, R., & Rück, C. (2017a). Heritability of hoarding symptoms across adolescence and young adulthood: A longitudinal twin study. PloS One, 12(6). https://doi.org/10.1371/JOURNAL.PONE.0179541

Ivanov, V. Z., Nordsletten, A., Mataix-Cols, D., Serlachius, E., Lichtenstein, P., Lundström, S., Magnusson, P. K. E., Kuja-Halkola, R., & Rück, C. (2017b). Heritability of hoarding symptoms across adolescence and young adulthood: A longitudinal twin study. PLOS ONE, 12(6), e0179541. https://doi.org/10.1371/JOURNAL.PONE.0179541

Lam, M., Awasthi, S., Watson, H. J., Goldstein, J., Panagiotaropoulou, G., Trubetskoy, V., Karlsson, R., Frei, O., Fan, C. C., De Witte, W., Mota, N. R., Mullins, N., Brügger, K., Hong Lee, S., Wray, N. R., Skarabis, N., Huang, H., Neale, B., Daly, M. J., … Ripke, S. (2020). RICOPILI: Rapid Imputation for COnsortias PIpeLIne. Bioinformatics, 36(3), 930–933. https://doi.org/10.1093/bioinformatics/btz633

Lee, J. J., Wedow, R., Okbay, A., Kong, E., Maghzian, O., Zacher, M., Nguyen-Viet, T. A., Bowers, P., Sidorenko, J., Karlsson Linnér, R., Fontana, M. A., Kundu, T., Lee, C., Li, H., Li, R., Royer, R., Timshel, P. N., Walters, R. K., Willoughby, E. A., … Turley, P. (2018). Gene discovery and polygenic prediction from a genome-wide association study of educational attainment in 1.1 million individuals. Nature Genetics, 50(8), 1112–1121. https://doi.org/10.1038/s41588-018-0147-3

Lee, P. H., Anttila, V., Won, H., Feng, Y. C. A., Rosenthal, J., Zhu, Z., Tucker-Drob, E. M., Nivard, M. G., Grotzinger, A. D., Posthuma, D., Wang, M. M. J., Yu, D., Stahl, E. A., Walters, R. K., Anney, R. J. L., Duncan, L. E., Ge, T., Adolfsson, R., Banaschewski, T., … Smoller, J. W. (2019). Genomic Relationships, Novel Loci, and Pleiotropic Mechanisms across Eight Psychiatric Disorders. Cell, 179(7), 1469-1482.e11. https://doi.org/10.1016/j.cell.2019.11.020

Lee, S. H., Wray, N. R., Goddard, M. E., & Visscher, P. M. (2011). Estimating missing heritability for disease from genome-wide association studies. Am J Hum Genet, 88(3), 294–305. https://doi.org/10.1016/j.ajhg.2011.02.002

Liang, K. Y., Wang, Y., Yin, Y. S., Grados, M., Fyer, A. J., Rauch, S., Murphy, D., McCracken, J., Rasmussen, S., Cullen, B., Hoehn-Saric, R., Greenberg, B., Pinto, A., Knowles, J., Piacentini, J., Pauls, D., Bienvenu, O., Riddle, M., Samuels, J., & Nestadt, G. (2008). Evidence for potential relationship between SLC1A1 and a putative genetic linkage region on chromosome 14q to obsessive-compulsive disorder with compulsive hoarding. American Journal of Medical Genetics, Part B: Neuropsychiatric Genetics, 147(6), 1000–1002. https://doi.org/10.1002/AJMG.B.30713

Lochner, C., Kinnear, C. J., Hemmings, S. M. J., Seller, C., Niehaus, D. J. H., Knowles, J. A., Daniels, W., Moolman-Smook, J. C., Seedat, S., & Stein, D. J. (2005). Hoarding in obsessive-compulsive disorder: Clinical and genetic correlates. Journal of Clinical Psychiatry, 66(9), 1155–1160. https://doi.org/10.4088/JCP.V66N0911

López-Solà, C., Fontenelle, L. F., Alonso, P., Cuadras, D., Foley, D. L., Pantelis, C., Pujol, J., Yücel, M., Cardoner, N., Soriano-Mas, C., Menchón, J. M., & Harrison, B. J. (2014). Prevalence and heritability of obsessive-compulsive spectrum and anxiety disorder symptoms: A survey of the Australian Twin Registry. American Journal of Medical Genetics. Part B, Neuropsychiatric Genetics_J_: The Official Publication of the International Society of Psychiatric Genetics, 165B(4), 314–325. https://doi.org/10.1002/AJMG.B.32233

Mathews, C. A., Delucchi, K., Cath, D. C., Willemsen, G., & Boomsma, D. I. (2014). Partitioning the etiology of hoarding and obsessive-compulsive symptoms. Psychological Medicine, 44(13), 2867–2876. https://doi.org/10.1017/S0033291714000269

Mathews, Carol A., Uhm, S., Chan, J., Gause, M., Franklin, J., Plumadore, J., Stark, S. J., Yu, W., Vigil, O., Salazar, M., Delucchi, K. L., & Vega, E. (2016). Treating Hoarding Disorder in a real-world setting: Results from the Mental Health Association of San Francisco. Psychiatry Research, 237, 331–338. https://doi.org/10.1016/J.PSYCHRES.2016.01.019

McCarthy, S., Das, S., Kretzschmar, W., Delaneau, O., Wood, A. R., Teumer, A., Kang, H. M.,Fuchsberger, C., Danecek, P., Sharp, K., Luo, Y., Sidore, C., Kwong, A., Timpson, N., Koskinen, S., Vrieze, S., Scott, L. J., Zhang, H., Mahajan, A., … Marchini, J. (2016). A reference panel of 64,976 haplotypes for genotype imputation. Nature Genetics 2016 48:10, 48(10), 1279–1283. https://doi.org/10.1038/ng.3643

Middeldorp, C. M., Hammerschlag, A. R., Ouwens, K. G., Groen-Blokhuis, M. M., St. Pourcain, B., Greven, C. U., Pappa, I., Tiesler, C. M. T., Ang, W., Nolte, I. M., Vilor-Tejedor, N., Bacelis, J., Ebejer, J. L., Zhao, H., Davies, G. E., Ehli, E. A., Evans, D. M., Fedko, I. O., Guxens, M., … Boomsma, D. I. (2016). A Genome-Wide Association Meta-Analysis of Attention-Deficit/Hyperactivity Disorder Symptoms in Population-Based Pediatric Cohorts. Journal of the American Academy of Child and Adolescent Psychiatry, 55(10), 896-905.e6. https://doi.org/10.1016/J.JAAC.2016.05.025

Nordsletten, A. E., Reichenberg, A., Hatch, S. L., Fernández De La Cruz, L., Pertusa, A., Hotopf, M., & Mataix-Cols, D. (2013). Epidemiology of hoarding disorder. The British Journal of Psychiatry_J_: The Journal of Mental Science, 203(6), 445–452. https://doi.org/10.1192/BJP.BP.113.130195

Park, L. S., Burton, C. L., Dupuis, A., Shan, J., Storch, E. A., Crosbie, J., Schachar, R. J., & Arnold, P. D. (2016). The Toronto Obsessive-Compulsive Scale: Psychometrics of a Dimensional Measure of Obsessive-Compulsive Traits. Journal of the American Academy of Child and Adolescent Psychiatry, 55(4), 310-318.e4. https://doi.org/10.1016/J.JAAC.2016.01.008

Perroud, N., Guipponi, M., Pertusa, A., Fullana, M. A., Iervolino, A. C., Cherkas, L., Spector, T., Collier, D., & Mataix-Cols, D. (2011). Genome-wide association study of hoarding traits. American Journal of Medical Genetics. Part B, Neuropsychiatric Genetics_J_: The Official Publication of the International Society of Psychiatric Genetics, 156(2), 240–242. https://doi.org/10.1002/AJMG.B.31152

Pertusa, A., Fullana, M. A., Singh, S., Alonso, P., Menchón, J. M., & Mataix-Cols, D. (2008). Compulsive hoarding: OCD symptom, distinct clinical syndrome, or both? The American Journal of Psychiatry, 165(10), 1289–1298. https://doi.org/10.1176/APPI.AJP.2008.07111730

Postlethwaite, A., Kellett, S., & Mataix-Cols, D. (2019). Prevalence of Hoarding Disorder: A systematic review and meta-analysis. Journal of Affective Disorders, 256, 309–316. https://doi.org/10.1016/J.JAD.2019.06.004

R core team. (2017). R: A language and environment for statistical computing. R Foundation for Statistical Computing, Vienna, Austria. URL https://Www.R-Project.Org/.

Rosseel, Y. (2012). lavaan: An R Package for Structural Equation Modeling. Journal of Statistical Software, 48, 1–36. https://doi.org/10.18637/JSS.V048.I02

Samuels, J., Joseph Bienvenu, O., Riddle, M. A., Cullen, B. A. M., Grados, M. A., Liang, K. Y., Hoehn-Saric, R., & Nestadt, G. (2002). Hoarding in obsessive compulsive disorder: results from a case-control study. Behaviour Research and Therapy, 40(5), 517–528. https://doi.org/10.1016/S0005-7967(01)00026-2

Samuels, J., Yin, Y. S., Grados, M. A., Willour, V. L., Bienvenu, O. J., Greenberg, B. D., Knowles, J. A., McCracken, J. T., Rauch, S. L., Murphy, D. L., Wang, Y., Pinto, A., Fyer, A. J., Piacentini, J., Pauls, D. L., Cullen, B., Rasmussen, S. A., Hoehn-Saric, R., Valle, D., … Nestadt, G. (2007). Significant linkage to compulsive hoarding on chromosome 14 in families with obsessive-compulsive disorder: results from the OCD Collaborative Genetics Study. The American Journal of Psychiatry, 164(3), 493–499. https://doi.org/10.1176/AJP.2007.164.3.493

Sinopoli, V. M., Erdman, L., Burton, C. L., Park, L. S., Dupuis, A., Shan, J., Goodale, T., Shaheen, S. M., Crosbie, J., Schachar, R. J., & Arnold, P. D. (2020). Serotonin system genes and hoarding with and without other obsessive–compulsive traits in a population-based, pediatric sample: A genetic association study. Depression and Anxiety, 37(8), 760–770. https://doi.org/10.1002/DA.22996

Steketee, G., Kelley, A. A., Wernick, J. A., Muroff, J., Frost, R. O., & Tolin, D. F. (2015). FAMILIAL PATTERNS OF HOARDING SYMPTOMS. Depression and Anxiety, 32(10), 728–736. https://doi.org/10.1002/DA.22393

Sudlow, C., Gallacher, J., Allen, N., Beral, V., Burton, P., Danesh, J., Downey, P., Elliott, P., Green, J., Landray, M., Liu, B., Matthews, P., Ong, G., Pell, J., Silman, A., Young, A., Sprosen, T., Peakman, T., & Collins, R. (2015). UK Biobank: An Open Access Resource for Identifying the Causes of a Wide Range of Complex Diseases of Middle and Old Age. PLOS Medicine, 12(3), e1001779. https://doi.org/10.1371/JOURNAL.PMED.1001779

Taylor, S., Jang, K. L., & Asmundson, G. J. G. (2010). Etiology of obsessions and compulsions: a behavioral-genetic analysis. Journal of Abnormal Psychology, 119(4), 672–682. https://doi.org/10.1037/A0021132

The 1000 Genomes Project Consortium. (2015). A global reference for human genetic variation. Nature, 526(7571), 68–74. https://doi.org/10.1038/nature15393

Timpano, K. R., Broman-Fulks, J. J., Glaesmer, H., Exner, C., Rief, W., Olatunji, B. O., Keough, M. E., Riccardi, C. J., Brähler, E., Wilhelm, S., & Schmidt, N. B. (2013). A taxometric exploration of the latent structure of hoarding. Psychological Assessment, 25(1), 194–203. https://doi.org/10.1037/A0029966

Timpano, K. R., Schmidt, N. B., Wheaton, M. G., Wendland, J. R., & Murphy, D. L. (2011). Consideration of the BDNF gene in relation to two phenotypes: hoarding and obesity. Journal of Abnormal Psychology, 120(3), 700–707. https://doi.org/10.1037/A0024159

Tolin, D. F., Frost, R. O., & Steketee, G. (2010). A brief interview for assessing compulsive hoarding: The Hoarding Rating Scale-Interview. Psychiatry Research, 178(1), 147. https://doi.org/10.1016/J.PSYCHRES.2009.05.001

Tolin, D. F., Frost, R. O., Steketee, G., & Fitch, K. E. (2008). Family burden of compulsive hoarding: Results of an internet survey. Behaviour Research and Therapy, 46(3), 334–344. https://doi.org/10.1016/J.BRAT.2007.12.008

Tolin, D. F., & Villavicencio, A. (2011). Inattention, but not OCD, predicts the core features of hoarding disorder. Behaviour Research and Therapy, 49(2), 120–125. https://doi.org/10.1016/J.BRAT.2010.12.002

Watanabe, K., Stringer, S., Frei, O., Umićević Mirkov, M., de Leeuw, C., Polderman, T. J. C., van der Sluis, S., Andreassen, O. A., Neale, B. M., & Posthuma, D. (2019). A global overview of pleiotropy and genetic architecture in complex traits. Nature Genetics, 51(9), 1339–1348. https://doi.org/10.1038/s41588-019-0481-0

Watanabe, K., Taskesen, E., Van Bochoven, A., & Posthuma, D. (2017). Functional mapping and annotation of genetic associations with FUMA. Nat Commu, 8(1), 1–10. https://doi.org/10.1038/s41467-017-01261-5

Wendland, J. R., Moya, P. R., Timpano, K. R., Anavitarte, A. P., Kruse, M. R., Wheaton, M. G., Ren-Patterson, R. F., & Murphy, D. L. (2009). A Haplotype Containing Quantitative Trait Loci for SLC1A1 Gene Expression and Its Association With Obsessive-Compulsive Disorder. Archives of General Psychiatry, 66(4), 408–416. https://doi.org/10.1001/ARCHGENPSYCHIATRY.2009.6

Willer, C. J., Li, Y., & Abecasis, G. R. (2010). METAL: Fast and efficient meta-analysis of genomewide association scans. Bioinformatics, 26(17), 2190–2191. https://doi.org/10.1093/bioinformatics/btq340

Wood, A. R., Esko, T., Yang, J., Vedantam, S., Pers, T. H., Gustafsson, S., Chu, A. Y., Estrada, K., Luan, J., Kutalik, Z., Amin, N., Buchkovich, M. L., Croteau-Chonka, D. C., Day, F. R., Duan, Y., Fall, T., Fehrmann, R., Ferreira, T., Jackson, A. U., … Frayling, T. M. (2014). Defining the role of common variation in the genomic and biological architecture of adult human height. Nature Genetics, 46(11), 1173–1186. https://doi.org/10.1038/NG.3097

World Health Organization. (2019). ICD-11: International classification of diseases (11th revision). Retrieved from https://Icd.Who.Int/.

Wunderle, V. M., Critcher, R., Ashworth, A., & Goodfellow, P. N. (1996). Cloning and Characterization ofSOX5,a New Member of the HumanSOXGene Family. Genomics, 36(2), 354–358. https://doi.org/10.1006/GENO.1996.0474

Yang, J., Lee, S. H., Goddard, M. E., & Visscher, P. M. (2011). GCTA: a tool for genome-wide complex trait analysis. American Journal of Human Genetics, 88(1), 76–82. https://doi.org/10.1016/J.AJHG.2010.11.011

Zhang, H., Leckman, J. F., Pauls, D. L., Tsai, C. P., Kidd, K. K., & Rosario Campos, M. (2002). Genomewide Scan of Hoarding in Sib Pairs in Which Both Sibs Have Gilles de la Tourette Syndrome. American Journal of Human Genetics, 70(4), 896. https://doi.org/10.1086/339520

Zheng, J., Erzurumluoglu, A. M., Elsworth, B. L., Kemp, J. P., Howe, L., Haycock, P. C., Hemani, G., Tansey, K., Laurin, C., Pourcain, B. S., Warrington, N. M., Finucane, H. K., Price, A. L., Bulik-Sullivan, B. K., Anttila, V., Paternoster, L., Gaunt, T. R., Evans, D. M., & Neale, B. M. (2017). LD Hub: A centralized database and web interface to perform LD score regression that maximizes the potential of summary level GWAS data for SNP heritability and genetic correlation analysis. Bioinformatics, 33(2), 272–279. https://doi.org/10.1093/bioinformatics/btw613

Zilhao, N. R., Smit, D. J., Boomsma, D. I., & Cath, D. C. (2016). Cross-disorder genetic analysis of tic disorders, obsessive-compulsive, and hoarding symptoms. Frontiers in Psychiatry, 7(JUN). https://doi.org/10.3389/fpsyt.2016.00120

