## Supplementary Tables for "Meta-Analysis of Genome-Wide Association Studies of Hoarding Symptoms in 27 537 Individuals"

**Supplementary Table S1:** Overview of Hoarding Rating Scale Self-Report (HRS-SR) questions in STR, NTR, and TwinsUK and the hoarding items of the Toronto Obsessive Compulsive Scale (TOCS) used in SfS.

| | STR | NTR | TwinsUK | SfS |
|---|---|---|---|---|
| **Questionnaire** | HRS-SR | HRS-SR | HRS-SR | TOCS |
| **Language** | Swedish | Dutch | English | English |
| **Responder** | Self-reported | Self-reported | Self-reported | Self-reported, parent-reported |
| **Time Period** | | | Retrospective over previous year | |
| **Possible range of answers** | 1-5 | 0-8 | 0-8 | |
| **item 1** | To what extent do you have difficulty discarding (or recycling, selling, giving away) ordinary things that other people would get rid of? (Original: Hur svårt har du att slänga vardagliga saker som andra lätt gör sig av med?) | Because of the clutter or number of possessions, how difficult is it for you to use the rooms in your home? | In the last year… Because of the clutter or number of possessions, how difficult is it for you to use the rooms in your home? | Collect useless objects |
| **item 2** | To what extent do you currently have a problem with collecting free things or buying more things than you need or can use or can afford? (Original: Hur stora problem har du med att du samlar på saker som är gratis eller att du köper mer saker än du behöver eller har råd med?) | To what extent do you have difficulty discarding (or recycling, selling, giving away) ordinary things that other people would get rid of? | In the last year…To what extent do you have difficulty discarding (or recycling, selling, giving away) ordinary things that other people would get rid of? | Have difficulty throwing things away |
| **item 3** | Because of the clutter or number of possessions, how difficult is it for you to use the rooms in your home? (Original: Hur svårt är det för dig att använda ditt rum och/eller dina utrymmen på grund av röran eller mängden saker?) | To what extent do you currently have a problem with collecting free things or buying more things than you need or can use or can afford? | In the last year…To what extent do you currently have a problem with collecting free things or buying more things than you need or can use or can afford? | - |
| **item 4** | To what extent do you experience emotional distress because of clutter, difficulty discarding or problems with buying or acquiring things? (Original: Hur känslomässigt upprörd blir du för att det är rörigt, eller dina svårigheter med att slänga, köpa eller skaffa saker?) | To what extent do you feel impaired (for example in your daily routine, job/eductaion, social activities) because of clutter, difficulty throwing things away, or problems with buying or acquiring things? | In the last year…To what extent do you experience impairment in your life (daily routine, job/school, social activities, family activities, fmancial difficulties) because of clutter, difficulty discarding, or problems with buying or acquiring things? | - |

**Supplementary Table S2:** LD-independent genomic regions with p < 1x10-5 in the HS meta-analysis and their associated genes. Listing for each single-nucleotide polymorphism (SNP) the respective chromosome (CHR), hg19 basepair-position (BP), association p-value (P), beta value from the association for allele 1 (Beta), standard error of the Beta (SE), effect allele and reference allele (A1/A2), frequency of allele 1 (FRQ), imutation quality score (INFO), number of individuals per SNP (N), direction of effect for each study (in the order STR-CatssGSA, STR-CatssPC, SfS, TwinsUK, NTR, STR-Yatss), a list of all variants with LD-r2 > 0.1 to the index SNP (friends(.1).p0.001), in brackets LD-r2 and distance in kb sorted by LD-r²; left and right margin of region (defined by LD friends) (range.left, range.right); span(kb)=right margin - left margin (in kb); same as before but with LD-r2 of 0.6 (friends(.6).p0.001, range.left.6, range.right.6, and span.6(kb)); list of entries in NHGRI GWAS catalogue among entries in column friends(.6) (gwas_catalog_span.6); and list of genes within the region of friends.6 (±50 kb), in brackets distance to index SNP in kb (genes.6.50kb(dist2index)).

| SNP | CHR | BP | P | Beta | SE | A1A2 | FRQ | INFO | N | Dir | LD-friends(0.1).p0.001 | range.left | range.right | span(kb) | LD-friends(0.6). | range.left.6 | range.right.6 | span.6(kb) | gwas_catalog_span.6 | genes.6.50kb(dist2index) | N.genes.6.50kb |
|---|---|---|---|---|---|---|---|---|---|---|---|---|---|---|---|---|---|---|---|---|---|
| rs117321479 | 12 | 23775967 | 1.36E-07 | 0.127901 | 0.0243 | G/A | 0.0277 | 0.987 | 20,812 | ----?- | rs117855589(0.998/-0.218),rs117847767(0.995/-1.44),rs41488244(0.981/-3.37),rs117715560(0.98/-7.83),rs118067810(0.979/-1.58),rs41349746(0.978/-6.48),rs80331093(0.944/4.85),rs80241699(0.792/-14.9),rs9971956(0.713/10.1),rs73279911(0.711/15.1),rs116968442(0.709/28.1),rs1002815(0.709/11.4),rs12297318(0.709/23.9),rs11047024(0.708/28.1),rs11047019(0.708/21.5),rs12306256(0.707/26.7),rs11047018(0.706/21.4),rs12319311(0.705/33.1),rs11047029(0.705/34),rs11047025(0.705/29.3),rs16926496(0.704/15.7),rs74071391(0.704/18.5),rs11047034(0.668/42.6) | 23761067 | 23818567 | 57.5 | rs117855589(0.9 | 23761067 | 23818567 | 57.5 | - | SOX5(0.0) | 1 |
| rs78426839 | 2 | 220170598 | 3.12E-07 | 0.1162 | 0.0227 | A/G | 0.0378 | 0.91 | 15,594 | ++?+?+ | rs34561714(0.806/22.1) | 220170598 | 220192698 | 22.1 | rs34561714(0.80 | 220170598 | 220192698 | 22.1 | /(0.806)rs34561714:Heigh | TUBA4B(0.0),DNAJB2(0.0),PT | 6 |
| rs7567224 | 2 | 125107948 | 7.70E-07 | -0.0349021 | 0.0071 | C/T | 0.411 | 0.994 | 27,651 | ++++++ | rs56405601(0.742/-3.01),rs2420840(0.718/21.4),rs62171019(0.71/5.04),rs72839455(0.619/-3.09),rs7582767(0.587/0.416),rs7609391(0.587/0.504),rs59503656(0.58/5.74),rs7581635(0.579/12.6),rs62171016(0.514/2.33),rs4848240(0.496/25.3),rs11123037(0.365/23.1),rs10496629(0.327/26.9),rs7578650(0.301/5.03),rs779999(0.292/0.077),rs780045(0.289/-5.38),rs780048(0.289/-13.8),rs780046(0.289/-6.59),rs780000(0.288/-2.31),rs796641(0.279/0.243),rs811902(0.268/-4.72),rs780057(0.266/23.5),rs720477(0.266/19.6),rs780059(0.265/25.6),rs779981(0.265/18),rs780060(0.262/27.4) | 125094148 | 125135348 | 41.2 | rs56405601(0.74 | 125104858 | 125129348 | 24.49 | - | CNTNAP5(0.0) | 1 |
| rs72927972 | 18 | 51528445 | 9.09E-07 | 0.0461963 | 0.0094 | A/C | 0.165 | 0.993 | 27,651 | -+++++ | rs72927946(0.997/-4.14),rs72926096(0.973/-11.6),rs72927915(0.973/-8.7),rs72926051(0.965/-18.4),rs72924148(0.963/-35.3),rs1498146(0.963/-43),rs72922092(0.962/-44.1),rs11660673(0.962/-41.6),rs142123239(0.957/-22.3),rs74652159(0.956/-62.1),rs72920351(0.956/-62.4),rs72920352(0.956/-62.3),rs72920339(0.956/-63.3),rs72929977(0.952/13.6),rs11659750(0.937/-6.26),rs72931916(0.937/21.6),rs79689252(0.893/-21),rs11661683(0.89/-23.4),rs75520509(0.88/-61.9),rs11659777(0.869/-72.7),rs17536164(0.658/-7.26),rs9304456(0.655/-7.82),rs28451198(0.654/-8.2),rs4502292(0.646/-16.1),rs28630974(0.644/-8.57),rs9955392(0.643/-12.2),rs16957645(0.643/-11.4),rs9955809(0.643/-11.6),rs4260126(0.643/-15.8),rs9962069(0.643/-10.1),rs7242057(0.643/-15.9),rs2133095(0.643/-13.7),rs76996980(0.643/-11.5),rs7236457(0.643/-10.5),rs7235741(0.643/-10.2),rs9946008(0.643/-14.6),rs4453581(0.643/-9.41),rs9951640(0.643/-16.8),rs7241181(0.643/-9.23),rs8092211(0.643/-16.9),rs34901938(0.643/-15.6),rs7231673(0.643/-10.6),rs16957651(0.643/-10.9),rs7239505(0.643/-13.4),rs16957628(0.643/-15.5),rs9947376(0.643/-11.5),rs7236245(0.643/-10.1),rs9955618(0.643/-11.7),rs9944731(0.643/-11.9),rs8091906(0.643/-17),rs7241585(0.643/-16.2),rs9958112(0.643/-11.5),rs16957626(0.643/-15.5),rs11664112(0.641/-17.1),rs72922067(0.641/-47.3),rs1039187(0.641/-51.1),rs11660620(0.641/-50.8),rs4041268(0.641/-49),rs11660649(0.641/-41.7),rs4299216(0.641/-51.9),rs11664547(0.641/-50),rs140719534(0.641/-23.1),rs72924158(0.641/- | 51450245 | 51670445 | 220.2 | rs72927946(0.99 | 51455745 | 51550045 | 94.3 | - | - | 0 |
| rs12713570 | 2 | 66875719 | 0.000001282 | -0.034105 | 0.007 | G/A | 0.433 | 0.997 | 27,651 | ++++++ | rs12713571(0.993/6.22),rs10490190(0.99/27.2),rs1974691(0.988/21.2),rs6720493(0.98/-31.9),rs12713569(0.873/-41.5),rs11695019(0.539/2.77),rs1558718(0.537/21.5),rs1303866(0.531/13.2),rs13006322(0.531/-5.85),rs55907104(0.528/4.65),rs34441555(0.528/28.1),rs13025385(0.528/29.9),rs17628190(0.528/27.9),rs6761229(0.521/-36.1),rs35751375(0.521/-15.7),rs2861111(0.519/-56.7),rs7602403(0.356/-43.1),rs11675077(0.356/27),rs57809212(0.338/-47),rs6754286(0.335/-50.3),rs61448585(0.331/-52.4),rs62146787(0.329/-46.1),rs6546236(0.32/-53.3),rs67054138(0.225/-8.67),rs55803117(0.225/-17.1),rs17032337(0.225/-10),rs9941653(0.225/21.5),rs140249857(0.225/-0.14),rs67981426(0.225/6.56),rs79707217(0.225/0.148),rs68044562(0.225/4.11),rs66923107(0.225/7.37),rs55830965(0.225/22.2),rs9941639(0.225/21),rs9309393(0.225/20.5),rs59788715(0.225/-1.25),rs55805041(0.225/3.4),rs75076546(0.224/-24.2),rs112315907(0.224/-27.1),rs67776981(0.224/-24.8),rs111521547(0.224/-15.7),rs7598188(0.224/-21.6),rs17032335(0.224/-11.8),rs6546239(0.224/-25.6),rs7573609(0.223/-26),rs6741487(0.156/-49.8),rs66533267(0.155/15.1),rs68170325(0.155/4.15),rs67294009(0.154/-10.1),rs192506056(0.154/-24.8),rs113982455(0.154/-10.9),rs67710556(0.154/5.75),rs2192962(0.154/-50.5),rs72808818(0.154/-12.9),rs60931071(0.154/7.44),rs113141186(0.154/-11.4),rs139535049(0.154/-18.8),rs181523914(0.153/-27.9),rs116704653(0.153/-27),rs4505509(0.153/-25.2),rs114128092(0.153/-27),rs60395529(0.153/-35.9),rs56311538(0.153/-22.1),rs55655907(0.152/-39.1) | 66819019 | 66905619 | 86.6 | rs12713571(0.99 | 66834219 | 66902919 | 68.7 | - | MEIS1(-24.7),LINC01798(0.0) | 3 |
| rs72838240 | 17 | 60701586 | 0.000001523 | -0.0917974 | 0.0191 | A/G | 0.0532 | 0.874 | 15,594 | +-?-?- | rs74999220(0.614/2.16),rs113512985(0.596/-53.3),rs112487967(0.591/-48.6),rs113427437(0.455/-196),rs112997374(0.116/-148),rs7223515(0.116/-136),rs9898829(0.116/-147),rs9675244(0.116/-136),rs72835233(0.115/-163),rs7406820(0.115/-148),rs1812692(0.115/-139),rs150830685(0.115/-145),rs112781672(0.114/-163),rs733025(0.11/-166) | 60505586 | 60703746 | 198.16 | rs74999220(0.61 | 60701586 | 60703746 | 2.16 | - | TLK2(0.0),MRC2(0.0) | 2 |

| | | | | | | | | | | | | | | | | | | | | | |
|---|---|---|---|---|---|---|---|---|---|---|---|---|---|---|---|---|---|---|---|---|---|
| rs140240461 | 7 | 85100327 | 0.00000226 | -0.154901 | 0.0327 | C/T | 0.0179 | 0.954 | 15,594 | ++?+?+ | rs117644446(0.899/-43),rs118119665(0.758/-56.3),rs117025998(0.747/58.9),rs117830687(0.728/137),rs10228353(0.423/-40.4),rs77217600(0.423/-19.3),rs11983565(0.423/-42.3),rs111935481(0.423/-35.9),rs76225781(0.422/-14.4),rs73389571(0.422/-13.9),rs74297487(0.421/-33.7),rs17159936(0.42/-29.3),rs10215813(0.42/-29.5),rs11979144(0.392/-51.8),rs2195554(0.387/-31.6),rs1546759(0.387/-28.5),rs2498551(0.383/-20.5),rs2463664(0.383/-20.4),rs2463665(0.383/-20.4),rs1403790(0.383/-13.1),rs2435279(0.382/-44.7),rs1608484(0.381/-44.3),rs1114589(0.381/-43.5),rs1589706(0.381/-43.9) | 85044027 | 85237327 | 193.3 | rs117644446(0.8 | 85044027 | 85237327 | 193.3 | - | LINC00972(0.0) | 1 |
| rs7534613 | 1 | 58971420 | 0.0000028 | -0.155497 | 0.0332 | C/T | 0.0176 | 0.919 | 15,594 | -+?+?+ | - | 58971420 | 58971420 | 0 | - | 58971420 | 58971420 | 0 | - | OMA1(0.0) | 1 |
| rs62467945 | 7 | 114508802 | 0.000003526 | 0.0416984 | 0.009 | T/A | 0.223 | 0.896 | 22,433 | --?--- | rs6945846(0.805/-106),rs62467929(0.686/-68.1),rs1840660(0.364/-156),rs62467874(0.354/-381),rs2045293(0.319/-159),rs13312596(0.319/-157),rs10500039(0.319/-158),rs13246064(0.312/-161),rs13222712(0.312/-160),rs776472(0.285/-156),rs12705984(0.266/-171),rs17583919(0.238/70.6),rs12705975(0.23/-193),rs1005959(0.23/-206),rs12705974(0.23/-193),rs12705971(0.23/-199),rs12705982(0.23/-173),rs1005958(0.23/-206),rs12705973(0.229/-196),rs13226763(0.229/-187),rs12705981(0.229/-174),rs1378771(0.229/-204),rs60234044(0.229/-206),rs1456021(0.229/-181),rs12671330(0.229/-175),rs12705977(0.229/-177),rs4730637(0.229/-205),rs12705978(0.229/-174),rs4236599(0.228/-191),rs2396766(0.228/-191),rs12154339(0.227/-194),rs2396765(0.225/-204),rs10250103(0.222/-214),rs1563408(0.215/-206),rs12705979(0.215/-174),rs17291908(0.215/-187),rs60603579(0.117/-473) | 114035802 | 114579402 | 543.6 | rs6945846(0.805 | 114402802 | 114508802 | 106 | - | - | 0 |
| rs77954641 | 19 | 33789608 | 0.000003693 | -0.0475949 | 0.0103 | T/G | 0.134 | 0.98 | 27,651 | ------ | rs111330623(0.998/0.071),rs73024223(0.996/0.586),rs41476047(0.995/1.03),rs12691(0.993/1.52),rs2081045(0.993/-0.598),rs1049969(0.991/2.2),rs34529039(0.991/3.02),rs707656(0.991/1.92),rs8102513(0.99/-2.67),rs8101436(0.99/-2.57),rs34508287(0.989/4.88),rs16967952(0.989/5.63),rs56160221(0.987/-3.4),rs56287732(0.966/10.7),rs73024299(0.956/12.7),rs8103337(0.953/15.2),rs59048871(0.953/13.8),rs8100151(0.953/14.8),rs111859293(0.95/16.7),rs57971673(0.516/31.1),rs74649479(0.319/-2.86),rs41504144(0.317/6.8),rs75061909(0.317/7.09),rs115349677(0.317/7.56),rs79347999(0.316/11.2),rs117035602(0.316/8.87),rs9646647(0.31/16.4),rs75279684(0.31/13.9),rs139037735(0.31/15.6),rs75710158(0.31/13.9),rs60572033(0.308/21.5),rs115178743(0.308/19.8),rs117674966(0.308/19.5),rs116956259(0.308/21),rs78079567(0.239/23.4),rs74954553(0.239/23.3),rs76778925(0.239/24.9),rs75306861(0.238/27.8),rs77949641(0.238/26.7),rs77886265(0.238/30.7),rs116916534(0.238/27),rs145852769(0.238/29.3),rs117915898(0.238/29.4),rs79955346(0.238/27.6),rs75225891(0.238/26.5),rs76243478(0.233/31.9),rs142997057(0.233/32.9),rs137915523(0.233/34.2),rs151115814(0.233/32.9),rs140470768(0.233/32.2),rs117866287(0.233/35.1),rs140086796(0.233/32),rs10451465(0.232/22.2),rs143406390(0.232/34.3),rs74334153(0.23/31.2) | 33786208 | 33824708 | 38.5 | rs111330623(0.9 | 33786208 | 33806308 | 20.1 | - | CEBPA(0.0),CEBPA-DT(0.0) | 2 |
| rs77736718 | 3 | 26882546 | 0.000005198 | -0.112396 | 0.0247 | T/C | 0.0319 | 0.945 | 15,594 | --?-?- | rs12632297(0.583/20.5),rs73063372(0.139/-30.8),rs17288364(0.108/-121) | 26761546 | 26903046 | 141.5 | - | 26882546 | 26882546 | 0 | - | - | 0 |
| rs144539704 | 9 | 136235935 | 0.00000524 | -0.139699 | 0.0307 | C/T | 0.0206 | 0.934 | 15,594 | ++?+?+ | rs78711238(0.945/-27.5),rs117308290(0.484/-106),rs117509822(0.472/4.97),rs11791989(0.215/-57.6),rs13301309(0.209/-71.1),rs11793580(0.199/-67),rs7020432(0.198/-70.2) | 136129935 | 136240905 | 110.97 | rs78711238(0.94 | 136208435 | 136235935 | 27.5 | - | SURF6(0.0),MED22(0.0),RPL7 | 13 |
| rs226114 | 11 | 110664239 | 0.000005287 | 0.0318959 | 0.007 | C/T | 0.499 | 0.966 | 27,651 | ------ | rs226115(0.978/-0.15),rs226112(0.786/0.691),rs670586(0.549/4.42),rs226164(0.543/22.5),rs226150(0.526/26.2),rs6589176(0.365/-12.8) | 110651439 | 110690439 | 39 | rs226115(0.978/- | 110664089 | 110664930 | 0.841 | - | - | 0 |
| rs9976960 | 21 | 24618326 | 0.000005572 | -0.0349953 | 0.0077 | T/G | 0.288 | 0.986 | 27,651 | ------ | rs9979890(0.996/1.5),rs9980697(0.996/1.47),rs9976409(0.841/-9.05),rs1380323(0.839/3.96),rs2061457(0.838/-11.3),rs9305168(0.701/-46.7),rs9981639(0.701/-46),rs7280897(0.7/-47.5),rs2244982(0.7/-18.5),rs985149(0.7/-47.7),rs2827952(0.699/-26.4),rs2827958(0.699/-25),rs2827951(0.699/-26.5),rs1506011(0.698/-50),rs6516558(0.698/-30),rs9305171(0.697/-44.9),rs1153197(0.697/-39.7),rs1506010(0.697/-49.9),rs1573359(0.697/-30.4),rs922081(0.697/-38.5),rs1506012(0.697/-50.1),rs1506009(0.697/-49.8),rs4392338(0.697/-43.4),rs1153203(0.697/-34.7),rs2827950(0.696/-50.2),rs1870464(0.694/-45.4),rs1870463(0.694/-45.4),rs9305166(0.694/-47.2),rs9982946(0.694/-45.4),rs9305167(0.693/-47.1),rs10854271(0.67/-27.4),rs9680422(0.67/-28.1),rs1153199(0.662/-38.7),rs4816894(0.657/-7.6),rs1870470(0.656/-7.43),rs8127651(0.655/-54.1),rs7276428(0.65/-69.9),rs2827948(0.649/-50.3),rs2827983(0.638/-9.63),rs2827979(0.637/-15.8),rs2827965(0.637/-22),rs13046121(0.635/-14.3),rs1304747(0.635/-33.1),rs2408668(0.635/-42.9),rs2169040(0.594/-2.28),rs198053(0.591/-105),rs2827987(0.579/0.523),rs1903161(0.561/7.25),rs2827992(0.555/5.56),rs4816896(0.543/9.15),rs2827995(0.543/8.99),rs2827994(0.542/8.85),rs768176(0.537/9.44),rs1380324(0.536/9.79),rs2226715(0.423/10.7),rs1380322(0.364/3.37),rs8133976(0.363/0.058),rs985211(0.363/1.91),rs1459399(0.362/-5.15),rs8132371(0.362/-1.57),rs1459402(0.362/-9.11),rs9977321(0.361/-14.1),rs2408670(0.361/-13),rs7282152(0.361/-8.95),rs1459400(0.361/-5.24) | 24513326 | 24629026 | 115.7 | rs9979890(0.996 | 24548426 | 24622286 | 73.86 | - | - | 0 |
| rs4817234 | 21 | 15899085 | 0.00000666 | -0.0366951 | 0.0082 | A/G | 0.248 | 0.942 | 27,651 | ------ | rs9979080(0.943/3.2),rs4817235(0.888/0.138) | 15899085 | 15902285 | 3.2 | rs9979080(0.943 | 15899085 | 15902285 | 3.2 | - | SAMSN1(0.0) | 1 |
| rs12708476 | 15 | 63293959 | 0.000007488 | 0.0314018 | 0.007 | G/A | 0.468 | 0.98 | 27,651 | ------ | rs12911467(0.684/-0.778),rs12903994(0.683/-2.08),rs34642222(0.655/-3.38),rs12913459(0.617/-3.59),rs11852619(0.304/-8.12),rs62011025(0.297/-17),rs17751112(0.294/-23.6),rs13380382(0.272/-8.38),rs17823147(0.266/-40),rs4352001(0.259/-32.3),rs28707628(0.258/-46.6),rs4775571(0.258/-43.6),rs2414805(0.257/-46.5),rs12900263(0.256/-46.7),rs35317361(0.255/-43.6),rs36030007(0.254/-26.8),rs2130162(0.253/-31.1),rs17750687(0.253/-28.9),rs12708475(0.253/-28.8),rs11631647(0.145/-37.1),rs28742917(0.123/21.5) | 63247259 | 63315459 | 68.2 | rs12911467(0.68 | 63290369 | 63293959 | 3.59 | - | TPM1(0.0),TPM1-AS(0.0) | 2 |

| | | | | | | | | | | | | | | | | | | | | | |
|---|---|---|---|---|---|---|---|---|---|---|---|---|---|---|---|---|---|---|---|---|---|
| rs148351785 | 6 | 134127406 | 0.000007711 | -0.155205 | 0.0347 | A/C | 0.0155 | 0.95 | 17,369 | ---??- | rs146099425(0.998/15.7),rs142898776(0.998/6.99),rs140466384(0.998/6.94),rs116851966(0.996/8.72),rs139120436(0.996/-0.923),rs138381522(0.996/-3.51),rs143307852(0.99/-11.8),rs143142433(0.986/-20.3),rs148236974(0.984/-20),rs117728942(0.977/-12),rs143798372(0.973/-28.4) | 134099006 | 134143106 | 44.1 | rs146099425(0.9 | 134099006 | 134143106 | 44.1 | - | TARID(0.0),LINC01312(0.0) | 2 |
| rs75688327 | 15 | 47351247 | 0.000007769 | -0.0488962 | 0.0109 | A/G | 0.137 | 0.88 | 22,433 | --?--- | rs1912977(0.317/-92.1),rs72729727(0.317/-99.7),rs4775660(0.317/-90.5),rs112835629(0.317/-81.4),rs2175575(0.317/-85.1),rs72729715(0.313/-113),rs72729717(0.313/-110),rs72729718(0.313/-109),rs10519093(0.313/-110),rs17279867(0.313/-112),rs55957745(0.313/-103),rs72729726(0.312/-101),rs16958906(0.312/-116),rs35378533(0.311/-102),rs4774487(0.311/-116),rs968233(0.311/-101),rs17279366(0.309/-124),rs16958894(0.309/-124),rs72727802(0.308/-127),rs72729720(0.308/-107),rs58025235(0.308/-131),rs17345891(0.308/-103),rs72727794(0.308/-133),rs72729722(0.308/-106),rs17345773(0.308/-109),rs1355589(0.308/-105),rs72729706(0.308/-124),rs17280083(0.308/-106),rs60580630(0.308/-105),rs11854171(0.306/-68.3),rs11857329(0.306/-68.3),rs4775663(0.306/-66.5),rs55927419(0.305/-64.7),rs72729723(0.292/-104),rs72729724(0.291/-104),rs72729725(0.29/-104),rs72727775(0.288/-148),rs56300189(0.288/-147),rs11855029(0.288/-164),rs73390803(0.287/-149),rs12148286(0.287/-151),rs56166124(0.287/-148),rs12148629(0.287/-142),rs11070560(0.255/-94.3),rs1918662(0.189/-257),rs17321993(0.175/-321),rs17270239(0.151/-268),rs10519079(0.15/-273),rs17268753(0.15/-326),rs2413831(0.15/-246),rs2413828(0.15/-309),rs2413832(0.15/-245),rs12902918(0.15/-270),rs66691168(0.15/-303),rs67761126(0.149/-320),rs12902816(0.149/-284),rs11854431(0.145/-265),rs12902436(0.145/-270),rs12912198(0.145/-247),rs4517728(0.145/-270) | 47025247 | 47351247 | 326 | - | 47351247 | 47351247 | 0 | - | - | 0 |
| rs12410615 | 1 | 57231991 | 0.00000788 | -0.0380031 | 0.0085 | A/G | 0.218 | 0.988 | 27,651 | ------ | rs1342381(0.997/-9.61),rs12022804(0.997/-9.03),rs2298127(0.996/-10.1),rs12410059(0.996/-6.29),rs12405047(0.785/-6.28),rs1578677(0.785/-4.06) | 57221891 | 57231991 | 10.1 | rs1342381(0.997 | 57221891 | 57231991 | 10.1 | - | PRKAA2(-1.0),FYB2(0.0) | 2 |
| rs79786919 | 2 | 47994675 | 0.000008542 | 0.115398 | 0.0259 | T/C | 0.029 | 0.9 | 15,594 | -+?+?+ | rs114144926(0.663/17),rs78731390(0.656/13.6),rs75733466(0.17/358),rs12053194(0.133/342) | 47994675 | 48352675 | 358 | rs114144926(0.6 | 47994675 | 48011675 | 17 | - | MSH6(0.0),FBXO11(0.0) | 2 |
| rs79269825 | 16 | 62453410 | 0.000009247 | -0.138504 | 0.0312 | T/C | 0.0205 | 0.928 | 14,988 | ?-?-?- | rs16964607(0.825/-36.9),rs117935849(0.795/-24.2),rs7194212(0.494/-60.5),rs6498848(0.494/-60.6),rs7194729(0.494/-60.3),rs16964587(0.493/-62.9),rs16964590(0.492/-62.8) | 62390510 | 62453410 | 62.9 | rs16964607(0.82 | 62416510 | 62453410 | 36.9 | - | - | 0 |
| rs76612314 | 4 | 10209522 | 0.000009445 | 0.0733991 | 0.0166 | A/G | 0.0725 | 0.859 | 15,594 | ++?+?+ | rs76835013(0.884/151),rs150196557(0.514/61.2),rs141474550(0.307/149),rs149844708(0.201/-152) | 10057522 | 10360522 | 303 | rs76835013(0.88 | 10209522 | 10360522 | 151 | - | - | 0 |
| 9:25926170_C_( | 9 | 25926170 | 0.000009717 | 0.100198 | 0.0226 | G/C | 0.0386 | 0.966 | 15,594 | --?-?- | rs10491877(0.975/-13.3),rs10812323(0.966/-10.7),rs7856714(0.964/-8.26),rs10967112(0.961/-26.1),rs10812324(0.96/7.26),rs7028344(0.956/5.2),rs75159073(0.955/2.87),rs77022124(0.953/-4.92),rs7026829(0.949/-12.9),rs7038828(0.939/-32.4),rs150384310(0.561/-5.68),rs1219936(0.281/-24.5),rs666778(0.264/-2.91),rs1219932(0.234/-19),rs644506(0.221/1.03),rs702228(0.221/-3.33),rs504517(0.221/-2.44),rs1782572(0.22/-11.4),rs542788(0.219/2.33),rs1219931(0.217/-17.5),rs562342(0.217/-0.669),rs658858(0.216/2.98),rs836957(0.216/-26.3),rs861359(0.214/1.32),rs636294(0.207/-3.76) | 25893770 | 25933430 | 39.66 | rs10491877(0.97 | 25893770 | 25933430 | 39.66 | - | - | 0 |

**Supplementary Table S3**: Results from the PRS analysis for HS (Leave-one-out), obsessive-compulsive disorder (OCD), depressive disorder (DEP), schizophrenia (SCZ), autism-spectrum disorder (ASD), attention-deficit hyperactivity disorder (ADHD), and educational attainment (EA). Results are presented for every target dataset (STR (combination of the three STR datasets), NTR, TwinsUK, and SfS), across pre-selected p-value thresholds (P Threshold). Listed are the Beta, standard error (SE), Z-score (Z), and p-value (P) from the regression. For STR, NTR, and TwinsUK the effective N was determined based on the actual N (including family members) weighted by the ratio of the squared SEs from the GEE sandwich-corrected model and the naive model (no correction). For SfS the sample N is listed. Bonferroni-corrected significant p-values (<0.05/7=0.00714) are in bold.

| Discovery | Target | P Threshold | Neffective (target) | Beta | SE | Z | P |
|---|---|---|---|---|---|---|---|
| ***Hoarding Symptoms (Leave-one-out)*** | | | | | | | |
| **HS** | **STR** | 0.5 | 11052.18 | 0.00181 | 0.00705 | 0.25689 | 0.79726 |
| | **NTR** | | 5759.06 | -0.00096 | 0.01101 | -0.08741 | 0.93035 |
| | **TwinsUK** | | 3359.35 | 0.00618 | 0.01288 | 0.48005 | 0.63119 |
| | **SfS** | | 5218 | 0.00993 | 0.01374 | 0.72246 | 0.47005 |
| ***Cross-trait*** | | | | | | | |
| **OCD** | **STR** | 0.1 | 11131.88 | -0.00439 | 0.00714 | -0.61446 | 0.53891 |
| | **NTR** | | 5717.05 | 0.00677 | 0.01128 | 0.60057 | 0.54813 |
| | **TwinsUK** | | 3398.42 | 0.01034 | 0.01279 | 0.80845 | 0.41883 |
| | **SfS** | | 5218 | 0.00993 | 0.01374 | 0.72246 | 0.47005 |
| **DEP** | **STR** | 0.05 | 10720.69 | 0.02448 | 0.00718 | 3.4082 | **0.00065** |
| | **NTR** | | 6118.68 | 0.00915 | 0.01112 | 0.82286 | 0.41059 |
| | **TwinsUK** | | 3548.09 | -0.03323 | 0.01253 | -2.65233 | **0.00799** |
| | **SfS** | | 5218 | 0.027333 | 0.01378 | 1.9842 | **0.04729** |
| **SCZ** | **STR** | 0.1 | 10680.06 | 0.03533 | 0.00747 | 4.72848 | **1x10-06** |
| | **NTR** | | 6071.04 | 0.01336 | 0.01198 | 1.11494 | 0.26488 |
| | **TwinsUK** | | 3578.2 | 0.02807 | 0.01399 | 2.00715 | **0.04473** |
| | **SfS** | | 5218 | 0.0025 | 0.0141 | 0.17727 | 0.85931 |
| **ASD** | **STR** | 0.1 | 11332.74 | 0.01465 | 0.00697 | 2.10171 | **0.03558** |
| | **NTR** | | 5981.45 | -0.0023 | 0.01126 | -0.2046 | 0.83789 |
| | **TwinsUK** | | 3444.56 | 0.0202 | 0.01295 | 1.56053 | 0.11863 |
| | **SfS** | | 5218 | 0.0328 | 0.01416 | 2.31679 | **0.02055** |
| **ADHD** | **STR** | 0.1 | 11173.34 | 0.01874 | 0.00706 | 2.65613 | **0.0079** |
| | **NTR** | | 6098.19 | -0.00152 | 0.01111 | -0.13706 | 0.89098 |
| | **TwinsUK** | | 3448.24 | -0.00913 | 0.01288 | -0.70911 | 0.47826 |
| | **SfS** | | 5218 | -0.01176 | 0.01384 | -0.84986 | 0.39544 |
| **EA** | **STR** | 1 | 10962.42 | 0.00749 | 0.00707 | 1.05896 | 0.28962 |
| | **NTR** | | 5786.57 | 0.02905 | 0.01121 | 2.59051 | **0.00958** |
| | **TwinsUK** | | 3340.07 | 0.05878 | 0.01298 | 4.5304 | **6x10-06** |
| | **SfS** | | 5218 | 0.02837 | 0.01472 | 1.92701 | 0.05403 |

Supplementary Table S4: Results of sign-test analysis for STR, NTR, SfS, and TwinsUK for three p-value thresholds (P_TH) 1e-06, 1e-05, and 1e-04. In the column "Discovery" are the discovery cohorts, in the coliumns "Replication" the target datasets. Nsum denotes the number of genomic regions in the replication study at each p-value threshold for which results are present. Npos is the number of genomic regions (in the replication study) that have the same direction with respect to the discovery results. The ratio is npos/nsum and sign-test is the P-value associated with the sign test. A ratio above 0.5 indicates a positive signtest, while a ratio below 0.5 indicates more divergence in the signtest than convergence.

| | | **Replication** | | | | | | | | | | | | | | | |
|---|---|---|---|---|---|---|---|---|---|---|---|---|---|---|---|---|---|
| | | **STR** | | | | **NTR** | | | | **SfS** | | | | **TwinsUK** | | | |
| **Discovery** | **P_TH** | **npos** | **nsum** | **sign-test** | **ratio** | **npos** | **nsum** | **sign-test** | **ratio** | **npos** | **nsum** | **sign-test** | **ratio** | **npos** | **nsum** | **sign-test** | **ratio** |
| **STR** | **0.0001** | | | | | 54 | 123 | 0.92559188 | 0.44 | 53 | 109 | 0.64909441 | 0.49 | 90 | 183 | 0.61621868 | 0.49 |
| **STR** | **0.00001** | | | | | 7 | 20 | 0.94234085 | 0.35 | 10 | 18 | 0.40726471 | 0.56 | 12 | 29 | 0.86753455 | 0.41 |
| **STR** | **0.000001** | | | | | 1 | 2 | 0.75 | 0.5 | 1 | 2 | 0.75 | 0.5 | 0 | 3 | 1 | 0 |
| **NTR** | **0.0001** | 54 | 123 | 0.92559188 | 0.44 | | | | | 27 | 53 | 0.5 | 0.51 | 39 | 69 | 0.16777881 | 0.57 |
| **NTR** | **0.00001** | 7 | 20 | 0.94234085 | 0.35 | | | | | 1 | 5 | 0.96875 | 0.2 | 4 | 5 | 0.1875 | 0.8 |
| **NTR** | **0.000001** | 1 | 2 | 0.75 | 0.5 | | | | | 0 | 1 | 1 | 0 | 0 | 1 | 1 | 0 |
| **SfS** | **0.0001** | 31 | 76 | 0.957677 | 0.41 | 30 | 67 | 0.83578397 | 0.45 | | | | | 33 | 76 | 0.89663158 | 0.43 |
| **SfS** | **0.00001** | 4 | 12 | 0.92700195 | 0.33 | 6 | 12 | 0.61279297 | 0.5 | | | | | 5 | 12 | 0.80615234 | 0.42 |
| **SfS** | **0.000001** | 0 | 0 | 1 | NA | 0 | 0 | 1 | NA | | | | | 0 | 0 | 1 | NA |
| **TwinsUK** | **0.0001** | 68 | 138 | 0.60072773 | 0.49 | 46 | 85 | 0.25771287 | 0.54 | 42 | 80 | 0.36877715 | 0.53 | | | | |
| **TwinsUK** | **0.00001** | 8 | 17 | 0.68547058 | 0.47 | 6 | 10 | 0.37695313 | 0.6 | 5 | 11 | 0.72558594 | 0.45 | | | | |
| **TwinsUK** | **0.000001** | 1 | 2 | 0.75 | 0.5 | 1 | 2 | 0.75 | 0.5 | 0 | 1 | 1 | 0 | | | | |

**Supplementary Table S5:** Results of sign-test analyses for STR. In the upper half, the discovery datasets are STR-only leave one out (LOO) analyses, in the lower half of the table, the discovery datasets are meta-analyses of NTR, TwinsUK, and LOO STR. Target/replication datasets are the individual STR datasets separated by cohort/age groups (Catss15 mean age = 15.47 (SD = 0.36); Catss18 mean age= 18.56 (SD = 0.33); Catss24 mean age = 23.84 (SD = 0.32); Yatss mean age = 23.93 (SD = 1.78) to determine if there was any pronounced age-related effect in the STR data. Sign-tests were performed for three different p-value thresholds (P_TH) 1e-06, 1e-05, and 1e-04. Nsum denotes the number of genomic regions in the replication study at each p-value threshold for which results are present. Npos is the number of genomic regions (in the replication study) that have the same direction with respect to the discovery results. The ratio is npos/nsum and 'sign-test' the P-value associated with the sign test. A ratio above 0.5 indicates a positive signtest, while a ratio below 0.5 indicates more divergence in the signtest then convergence.

| Base | Target | P_TH | npos | nsum | sign-test | ratio |
|---|---|---|---|---|---|---|
| **STR only** | | | | | | |
| Catss18PC_Catss24PC_Yatss | STR_Catss15_PC | 0.0001 | 101 | 194 | 0.30769037 | 0.52 |
| Catss18PC_Catss24PC_Yatss | STR_Catss15_PC | 0.00001 | 11 | 19 | 0.32380295 | 0.58 |
| Catss18PC_Catss24PC_Yatss | STR_Catss15_PC | 0.000001 | 2 | 2 | 0.25 | 1 |
| Catss15PC_Catss24PC_Yatss | STR_Catss18_PC | 0.0001 | 99 | 191 | 0.33214799 | 0.52 |
| Catss15PC_Catss24PC_Yatss | STR_Catss18_PC | 0.00001 | 9 | 23 | 0.89498019 | 0.39 |
| Catss15PC_Catss24PC_Yatss | STR_Catss18_PC | 0.000001 | 0 | 5 | 1 | 0 |
| Catss15PC_Catss18PC_Yatss | STR_Catss24_PC | 0.0001 | 86 | 179 | 0.72500228 | 0.48 |
| Catss15PC_Catss18PC_Yatss | STR_Catss24_PC | 0.00001 | 13 | 22 | 0.26173353 | 0.59 |
| Catss15PC_Catss18PC_Yatss | STR_Catss24_PC | 0.000001 | 3 | 6 | 0.65625 | 0.5 |
| Catss15PC_Catss18PC_Catss24PC | STR_Yatss | 0.0001 | 100 | 190 | 0.25695482 | 0.53 |
| Catss15PC_Catss18PC_Catss24PC | STR_Yatss | 0.00001 | 18 | 29 | 0.13246545 | 0.62 |
| Catss15PC_Catss18PC_Catss24PC | STR_Yatss | 0.000001 | 0 | 3 | 1 | 0 |
| **All cohorts (replicating separate STR)** | | | | | | |
| NTR_TwinsUK_STRCatss18PC_Catss24PC_Yatss | STR_Catss15PC | 0.0001 | 85 | 155 | 0.13036604 | 0.55 |
| NTR_TwinsUK_STRCatss18PC_Catss24PC_Yatss | STR_Catss15PC | 0.00001 | 8 | 13 | 0.29052734 | 0.62 |
| NTR_TwinsUK_STRCatss18PC_Catss24PC_Yatss | STR_Catss15PC | 0.000001 | 2 | 3 | 0.5 | 0.67 |
| NTR_TwinsUK_STRCatss15PC_Catss24PC_Yatss | STR_Catss18PC | 0.0001 | 86 | 176 | 0.64681458 | 0.49 |
| NTR_TwinsUK_STRCatss15PC_Catss24PC_Yatss | STR_Catss18PC | 0.00001 | 8 | 17 | 0.68547058 | 0.47 |
| NTR_TwinsUK_STRCatss15PC_Catss24PC_Yatss | STR_Catss18PC | 0.000001 | 0 | 1 | 1 | 0 |
| NTR_TwinsUK_STRCatss15PC_Catss18PC_Yatss | STR_Catss24PC | 0.0001 | 86 | 168 | 0.40852324 | 0.51 |
| NTR_TwinsUK_STRCatss15PC_Catss18PC_Yatss | STR_Catss24PC | 0.00001 | 12 | 20 | 0.25172234 | 0.6 |
| NTR_TwinsUK_STRCatss15PC_Catss18PC_Yatss | STR_Catss24PC | 0.000001 | 2 | 3 | 0.5 | 0.67 |
| NTR_TwinsUK_STRCatss15PC_Catss18PC_Catss24PC | STR_Yatss | 0.0001 | 84 | 154 | 0.14740676 | 0.55 |
| NTR_TwinsUK_STRCatss15PC_Catss18PC_Catss24PC | STR_Yatss | 0.00001 | 6 | 12 | 0.61279297 | 0.5 |
| NTR_TwinsUK_STRCatss15PC_Catss18PC_Catss24PC | STR_Yatss | 0.000001 | 3 | 4 | 0.3125 | 0.75 |

**Supplementary Table S6:** Results of sign-test analyses for NTR, separated into three age groups (NTR < 30 years, NTR 30-45 years, NTR > 45 years). The discovery datasets are the over-all STR dataset, as well as the STR sub-cohorts separated by age. This was done to determine if there were any pronounced age-related effect in the NTR data. Sign-tests were performed for four different p-value thresholds (P_TH) 5e-08, 1e-06, 1e-05, and 1e-04. Nsum denotes the number of genomic regions in the replication study at each p-value threshold for which results are present. Npos is the number of genomic regions (in the replication study) that have the same direction with respect to the discovery results. The ratio is npos/nsum and sign-test the P-value associated with the sign test. A ratio above 0.5 indicates a positive signtest, while a ratio below 0.5 indicates more divergence in the signtest then convergence.

| | | NTR < 30 | | | | NTR 30-45 | | | | NTR > 45 | | | |
|---|---|---|---|---|---|---|---|---|---|---|---|---|---|
| **Discovery** | **P_TH** | **npos** | **nsum** | **sign-test** | **ratio** | **npos** | **nsum** | **sign-test** | **ratio** | **npos** | **nsum** | **sign-test** | **ratio** |
| **TwinsUK** | **0.0001** | 42 | 83 | 0.5 | 0.51 | 50 | 85 | 0.0641982 | 0.59 | 44 | 86 | 0.45710582 | 0.51 |
| **TwinsUK** | **0.00001** | 6 | 10 | 0.37695313 | 0.6 | 6 | 10 | 0.37695313 | 0.6 | 6 | 11 | 0.5 | 0.55 |
| **TwinsUK** | **0.000001** | 1 | 2 | 0.75 | 0.5 | 1 | 2 | 0.75 | 0.5 | 1 | 2 | 0.75 | 0.5 |
| **STR** | **0.0001** | 61 | 125 | 0.63966374 | 0.49 | 57 | 123 | 0.81635163 | 0.46 | 55 | 123 | 0.89667645 | 0.45 |
| **STR** | **0.00001** | 11 | 20 | 0.41190147 | 0.55 | 10 | 20 | 0.58809853 | 0.5 | 8 | 20 | 0.86841202 | 0.4 |
| **STR** | **0.000001** | 2 | 2 | 0.25 | 1 | 0 | 2 | 1 | 0 | 1 | 2 | 0.75 | 0.5 |
| **Catss15** | **0.0001** | 46 | 88 | 0.3746645 | 0.52 | 50 | 89 | 0.14454804 | 0.56 | 37 | 89 | 0.9553396 | 0.42 |
| **Catss15** | **0.00001** | 3 | 9 | 0.91015625 | 0.33 | 6 | 9 | 0.25390625 | 0.67 | 3 | 9 | 0.91015625 | 0.33 |
| **Catss15** | **0.000001** | 0 | 0 | 1 | NA | 0 | 0 | 1 | NA | 0 | 0 | 1 | NA |
| **Catss18** | **0.0001** | 47 | 94 | 0.54103847 | 0.5 | 47 | 91 | 0.41704052 | 0.52 | 50 | 91 | 0.2009064 | 0.55 |
| **Catss18** | **0.00001** | 5 | 9 | 0.5 | 0.56 | 3 | 8 | 0.85546875 | 0.38 | 1 | 9 | 0.9980469 | 0.11 |
| **Catss18** | **0.000001** | 0 | 1 | 1 | 0 | 0 | 1 | 1 | 0 | 1 | 1 | 0.5 | 1 |
| **Catss24** | **0.0001** | 45 | 87 | 0.4151865 | 0.52 | 36 | 84 | 0.92217177 | 0.43 | 39 | 84 | 0.77739767 | 0.46 |
| **Catss24** | **0.00001** | 4 | 10 | 0.828125 | 0.4 | 4 | 10 | 0.828125 | 0.4 | 2 | 10 | 0.9892578 | 0.2 |
| **Catss24** | **0.000001** | 0 | 0 | 1 | NA | 0 | 0 | 1 | NA | 0 | 0 | 1 | NA |
| **Yatss** | **0.0001** | 54 | 130 | 0.9783724 | 0.42 | 65 | 130 | 0.53492233 | 0.5 | 71 | 129 | 0.14534393 | 0.55 |
| **Yatss** | **0.00001** | 11 | 18 | 0.24034119 | 0.61 | 10 | 18 | 0.40726471 | 0.56 | 11 | 18 | 0.24034119 | 0.61 |
| **Yatss** | **0.000001** | 2 | 2 | 0.25 | 1 | 1 | 2 | 0.75 | 0.5 | 1 | 2 | 0.75 | 0.5 |
