## Supplementary Information for "Meta-Analysis of Genome-Wide Association Studies of Hoarding Symptoms in 27 537 Individuals"

### Supplement Hoarding Symptom GWAS

1. **Methods** 1
1.1. Cohort descriptions 1
1.2. Hoarding phenotype and symptom measures 3
1.2.1. Distribution of hoarding items and hoarding sum-scores in the different cohorts 3
1.2.2. Distributions of standardized Hoarding sum-scores 9
1.3. Genotyping, quality control and imputation of individual cohorts 11
2. **Results** 16
2.1. Genome-wide association results 16
2.2. Compatibility between cohorts 21

### 1. Methods

#### 1.1. Cohort descriptions

##### *STR*

Individuals included in this study were monozygotic (MZ) or dizygotic (DZ) twins enrolled in the population-based *Swedish Twin Registry* (STR), who participated in one of its large-scale cohort studies, namely the *Child and Adolescent Twin Study in Sweden* (CATSS15, CATSS18, CATSS24), or the *Young Adult Twins in Sweden Study* (YATSS). CATSS is a prospective, longitudinal study of all twins born in Sweden since 1992, measured at age 15 (CATSS15), age 18 (CATSS18), and/or age 24 (CATSS24) (see Anckarsäter et al., 2011 for details). CATSS data collection was initiated in 2004. The YATSS survey took place in 2013, including twin-pairs born between May 1986 and June 1992. For both studies, collected data types include phenotypic and exposure data gathered via questionnaires and interviews, as well as genotypic data through saliva samples. DNA extraction from saliva started in 2009 when kits to extract saliva were sent to the participants after the completion of the online-based or telephone interviews. DNA extraction was then performed at the Karolinska Institutet biobank. One part of the CATSS samples was genotyped on the PsychChip genotyping array (N = 8598), another part was genotyped on the GSA genotyping array (N = 606). All samples from CATSS15, CATSS18, and CATS24 that were genotyped together on either the GSA chip or the PsychChip were also analyzed together in the GWAS analysis. In the following they are called STR-CATSS-GSA and STR-CATSS-PC, each including individuals from CATSS15, CATSS18, and CATSS24 (More details are described in the chapter "*1.3 Genotyping, quality control and imputation of individual cohorts*"). Also, see Zagai et al. (2019) for details of the STR, and Ivanov et al., (2017) for hoarding specific details of the cohorts. All participants gave their informed consent. The study was approved under the reference number 2018/2232-32.

##### *NTR*

Twins and their family members (parents, children, siblings) registered at the Netherlands Twin Register (NTR) participated. Every two to three years subjects 16 years and older receive self-report surveys that contain a variety of questionnaires related to health, personality, demographics, lifestyle and psychiatric disorders (Boomsma et al., 2002). Data on hoarding symptoms were available for 16,003 subjects in the survey from 2008. Of these, N=6,545 (66% female) had genotype data available and were of Dutch ancestry. Their mean age was 41.4 years (SD 15.2; age range 17–70 years). Informed consent was obtained from all participants. The study was approved by the Central Ethics Committee on Research Involving Human Subjects of the VU University Medical Centre, Amsterdam, an Institutional Review Board certified by the U.S. Office of Human Research Protections (IRB number IRB00002991 under Federal-wide Assurance - FWA00017598; IRB/institute codes, NTR 03-180).

##### *SfS*

Spit for Science (SfS) is a population-based cohort of youth and adolescents recruited at the Ontario Science Center (OSC) in Toronto, Canada. The SfS sample has been previously described elsewhere (Ameis et al., 2016; Crosbie et al., 2013). Briefly, the SfS subset analyzed here included 5,218 unrelated participants of European ancestry with complete demographic and questionnaire information collected at the OSC over a 16-month period between 2008 to 2009. Self-report ethnicity was confirmed using genetic data. Protocols for informed consent and assent (where applicable) were approved by the Research Ethics Board at the Hospital for Sick Children. All participants provided saliva samples for genotyping.

##### *TwinsUK*

Participants were monozygotic and dizygotic twins from the TwinsUK adult twin registry (www.twinsuk.ac.uk). The characteristics of the sample are described elsewhere (Spector & Williams, 2006). This registry data set consists of approximately 10,000 monozygotic and dizygotic twin pairs of European ancestry, from the United Kingdom, aged 16 and above. Recruitment was through a series of media advertisements that did not target individuals on the basis of a pre-existing disorder. The characteristics of the cohort have been shown to be comparable with age-matched population singletons in terms of disease-associated and lifestyle characteristics (Andrew et al., 2001). Zygosity status was initially obtained using the “Peas in the Pod” questionnaire (Sarna et al., 1978) and has been further validated using genome-wide genetic data. The Hoarding Rating Scale–Self-Report (HRS-SR) was sent to all active twins in the registry (N = 8,313) as part of a larger wave of data collection. Analyzable questionnaire data was returned by 5,022 twins. There was no distinction between responders and non-responders in terms of zygosity (53% and 50% monozygotic, respectively) and socioeconomic status (Index of Multiple Deprivation mean score (see https://www.gov.uk/government/collections/english-indices-of-deprivation), 3.7 [SD = 1.2] compared with 3.3 [SD = 1.3], respectively). However, non-responders were more likely to be male (56% compared with 36%; $\chi 2 = 189.9$, df = 1, $p < 0.01$) and younger (mean age, 46 years [SD = 14, range = 16–90] compared with 55 years [SD = 13; range = 17–86]; t = 28.8, df = 8,312, $p < 0.01$). Hence the full-size UKTwins Hoarding data set analysis consists of 2,053 twin pairs (125 monozygotic males, 65 dizygotic males, 971 monozygotic females, 857 dizygotic females, and 35 dizygotic twins of the opposite sex) and 916 singleton twins (73 monozygotic males, 56 dizygotic males, 383 monozygotic females, 316 dizygotic females, 34 dizygotic opposite-sex twins, and 54 dizygotic twins whose co-twin sex was unknown). The ethics committee of The Guys & St Thomas' Trust (GSTT) gave ethical approval for the work related to the TwinsUK study.

#### 1.2. Hoarding phenotype and symptom measures

##### *1.2.1. Distribution of hoarding items and hoarding sum-scores in the different cohorts*

##### *STR*

**Supplementary Figure S1:** Distributions of questionnaire scores in the Hoarding Rating Scale Self-Report (HRS-SR) across the STR-CATSS-GSA samples. A-D shows score distributions for each of the HRS-SR items, E shows the score distribution of the sum-score of item 1-4.

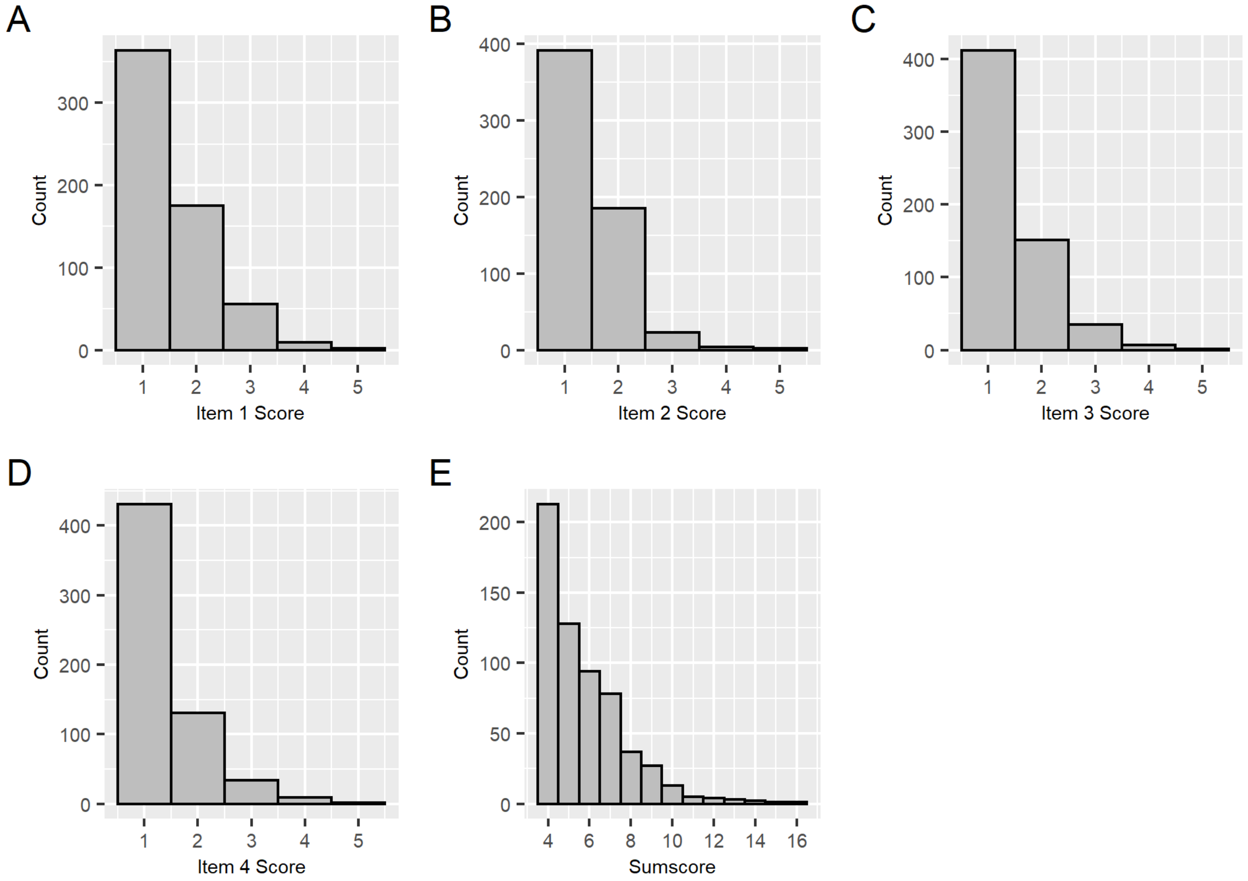

**Supplementary Figure S2:** Distributions of questionnaire scores in the Hoarding Rating Scale Self-Report (HRS-SR) across the STR-CATSS-PC samples. A-D shows score distributions for each of the HRS-SR items, E shows the score distribution of the sum-score of item 1-4.

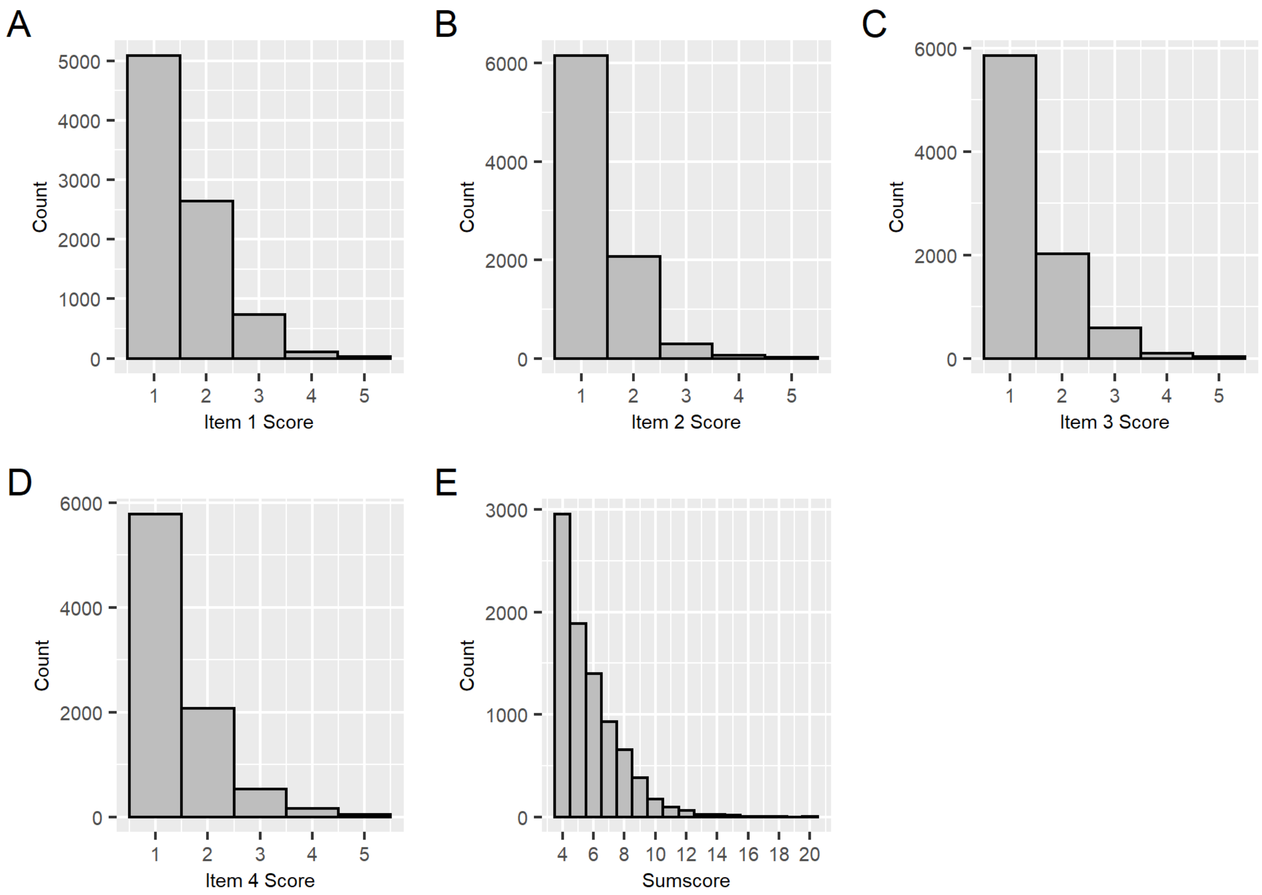

**Supplementary Figure S3:** Distributions of questionnaire scores in the Hoarding Rating Scale Self-Report (HRS-SR) across the YATSS samples in the STR. A-D shows score distributions for each of the HRS-SR items, E shows the score distribution of the sum-score of item 1-4.

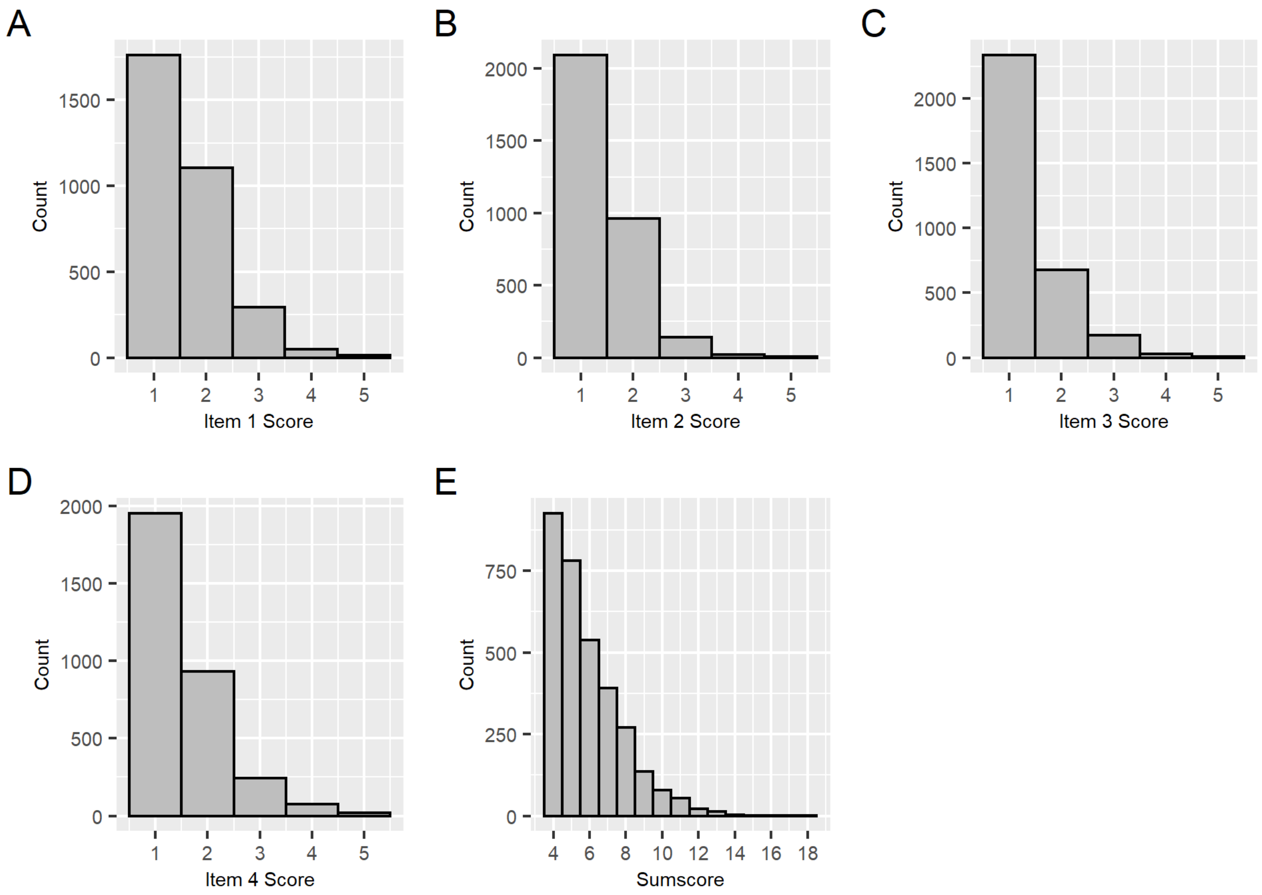

#### *NTR*

**Supplementary Figure S4:** Distributions of questionnaire scores in the Hoarding Rating Scale Self-Report (HRS-SR) across the NTR. A-D shows score distributions for each of the HRS-SR items, E shows the score distribution of the sum-score of item 1-4.

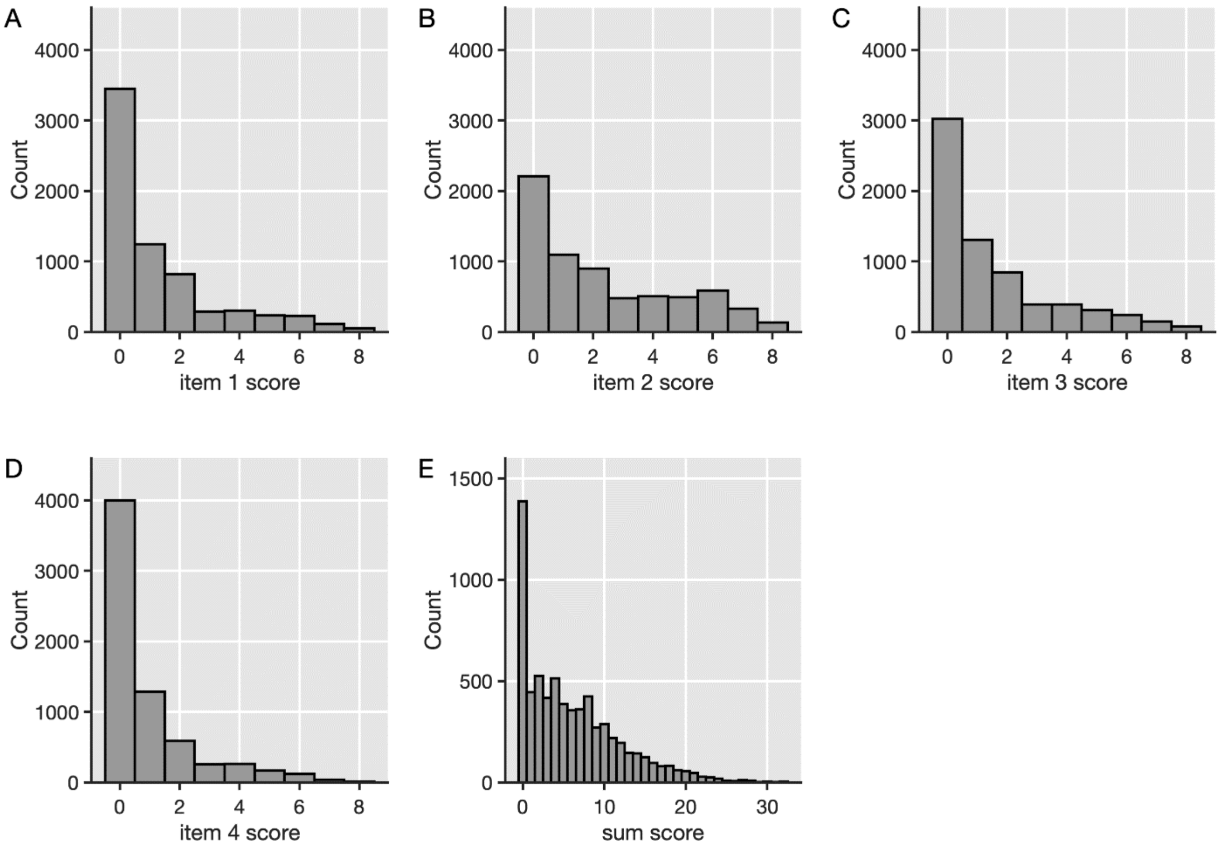

#### *SfS*

**Supplementary Figure S5.** Distributions of raw hoarding items 1 (A) and 2 (B) from the Toronto Obsessive-Compulsive Scale in Spit for Science. Total score (C) is the sum of the two raw items.

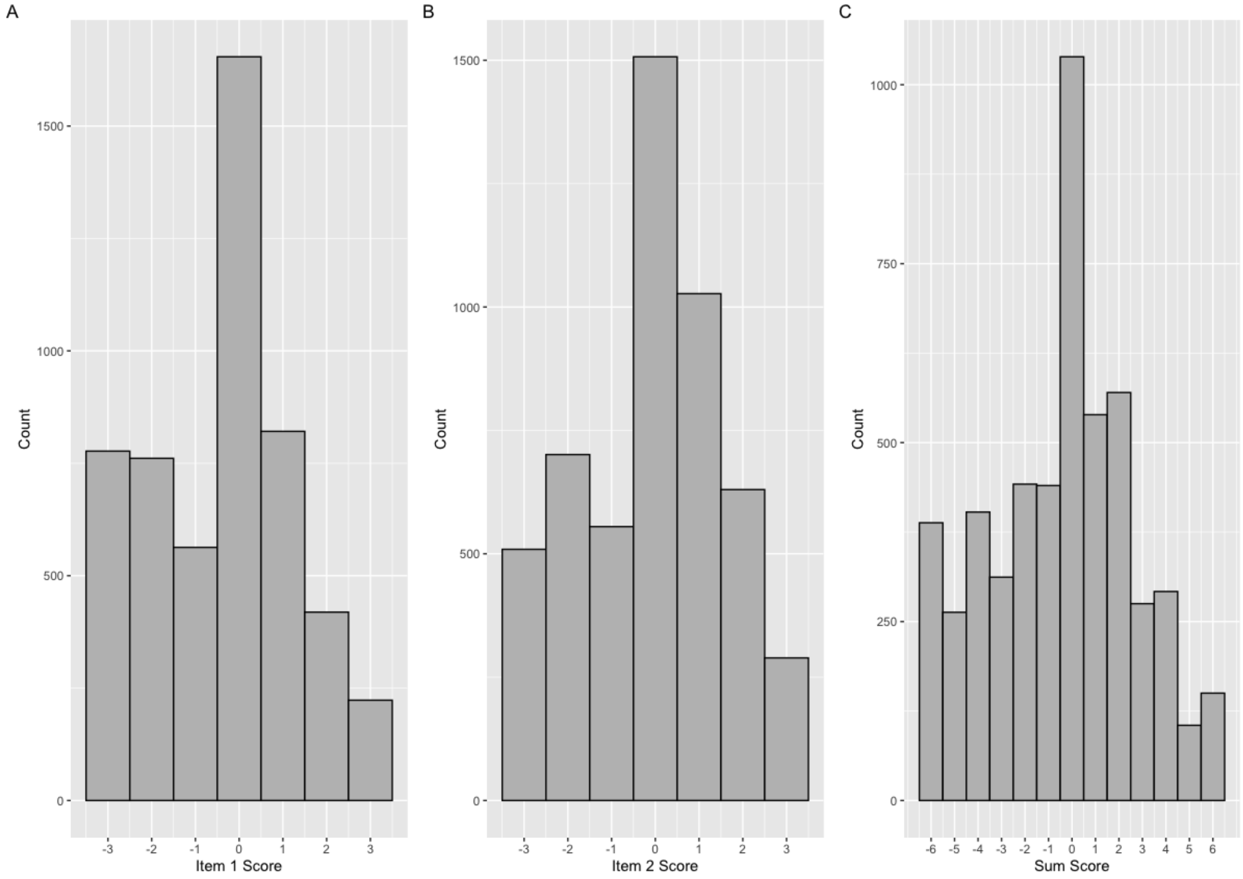

#### *TwinsUK*

**Supplementary Figure S6:** Distributions of questionnaire scores in the Hoarding Rating Scale Self-Report (HRS-SR) across TwinsUK. A-E shows score distributions for each of the HRS-SR items, F shows the score distribution of the sum-score of item 1-5.

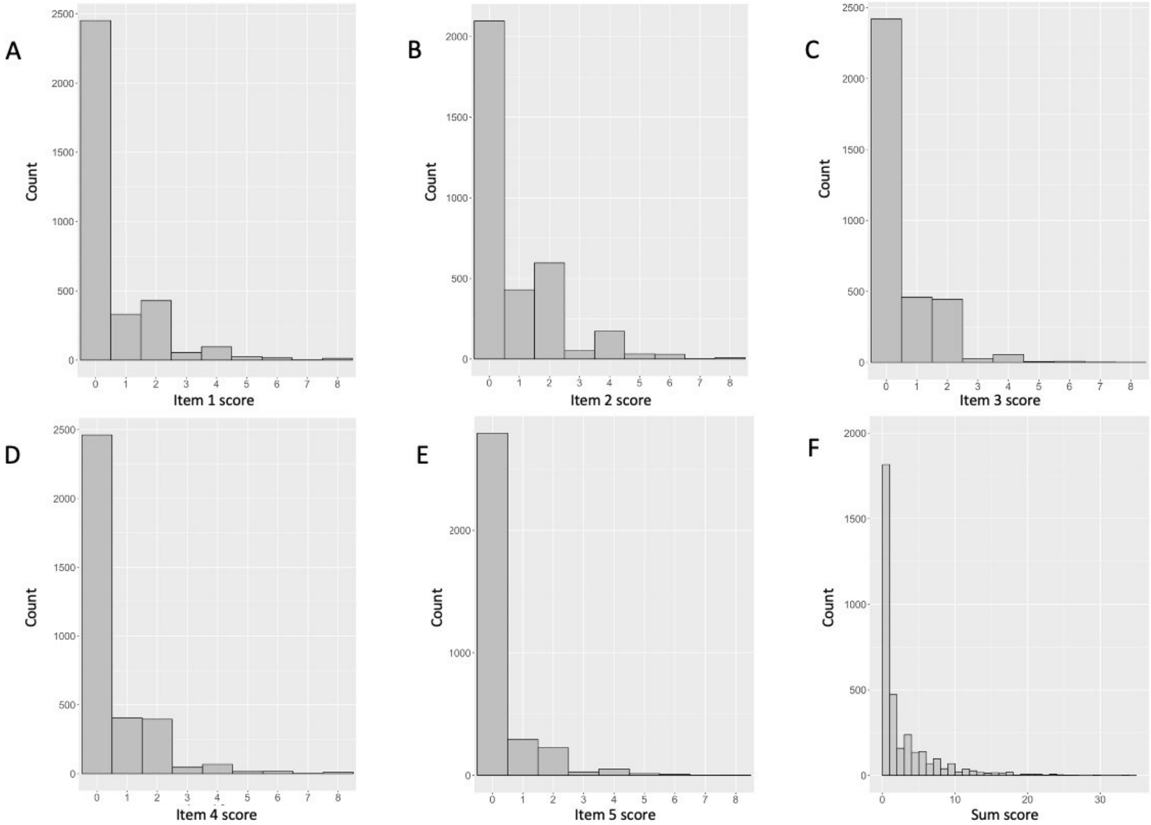

##### *1.2.2. Distributions of standardized Hoarding sum-scores*

##### *STR*

**Supplementary Figure S7:** Distributions of scores resulting from a one-factor model using latent factor analysis based on the four items of the HRS-SR separately for the CATSS-GSA (A), CATSS-PC (B), and YATSS (C) STR cohorts.

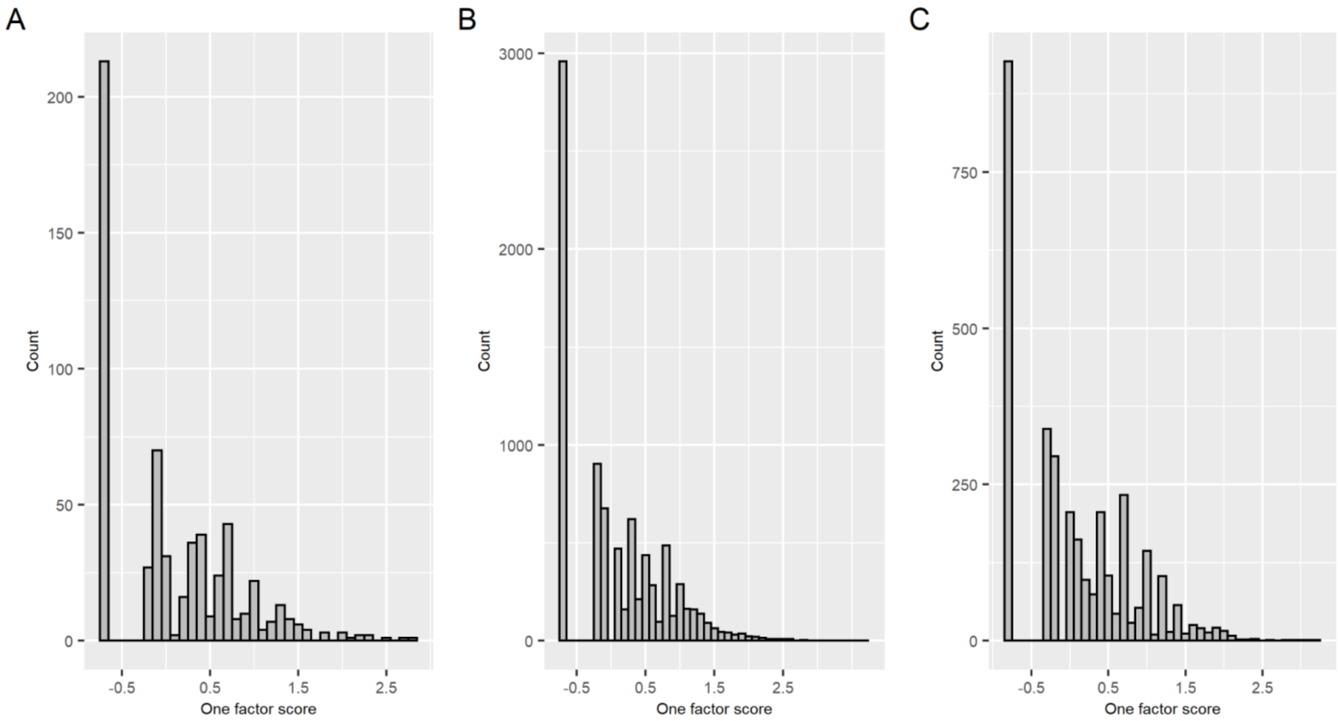


##### *NTR*

**Supplementary Figure S8:** Distribution of the score resulting from a one-factor model using latent factor analysis based on the four items of the HRS-SR in NTR.

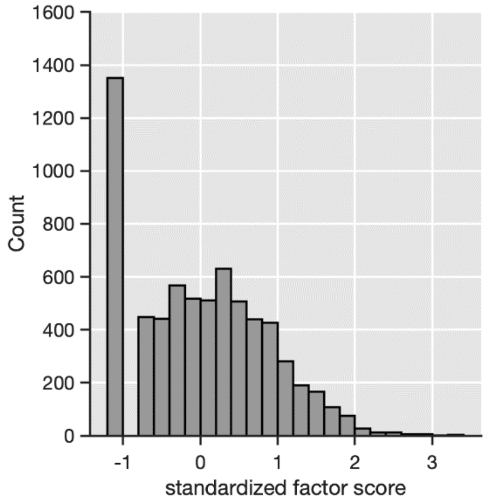

#### *TwinsUK*

**Supplementary Figure S9:** Distribution of the score resulting from a one-factor model using latent factor analysis based on the four items of the HRS-SR in NTR.

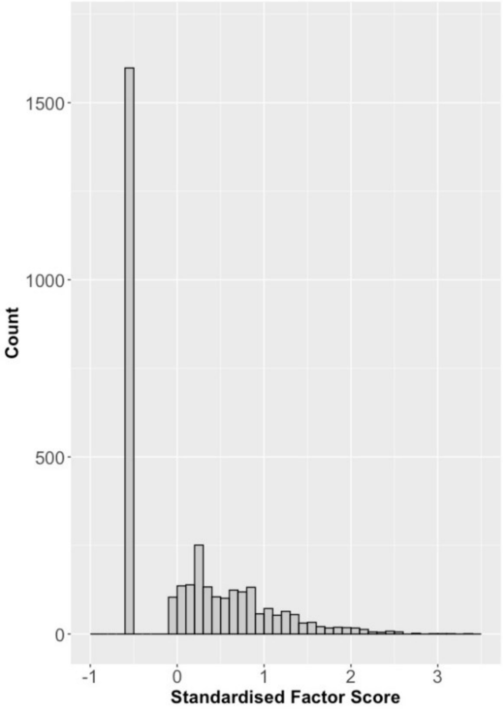


#### *SfS*

**Supplementary Figure S10:** Distribution of the standardized sum score of the two hoarding items from the Toronto Obsessive Compulsive Scale in Spit for Science.

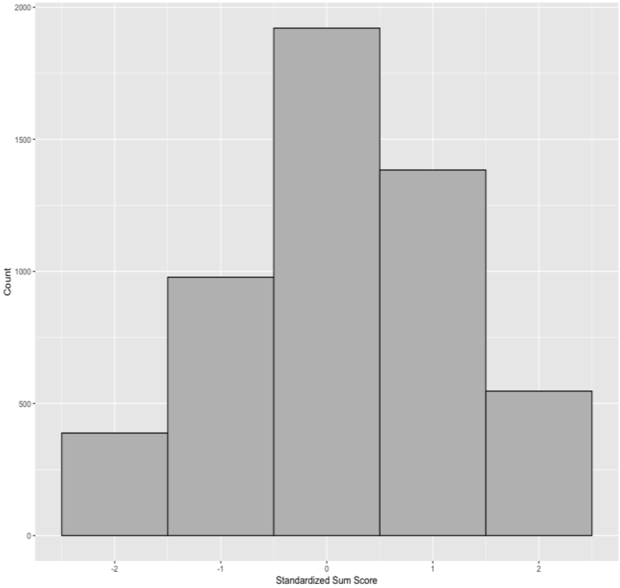

#### 1.3. Genotyping, quality control and imputation of individual cohorts

##### *STR*

The four STR cohorts (CATSS15, CATSS18, CATSS24, and YATSS) form three separate datasets (CATSS-GSA, CATSS-PC, and YATSS) that were each genotyped and imputed separately. CATSS-GSA and CATSS-PC each include samples from CATSS15, CATSS18, and CATSS24.

CATSS-PC samples were genotyped on the Illumina PsychChip array and were processed using the Ricopili (Lam et al., 2020) pipeline for quality control (QC). In a first round of QC, SNPs with a missingness higher than 0.05 (N = 4,477) were removed, followed by the exclusion of 141 samples due to either per-sample call rate < 0.98, excessive heterozygosity (FHET outside +/- 0.2), or sex mismatch. 146,755 out of 588,454 markers failed SNP QC due to either per-SNP call rate < 0.98, Hardy-Weinberg disequilibrium ($P < 1x10^{-6}$ in MZ twins and $P < 1x10^{-10}$ in DZ twins), or difference in call rate between MZ twins and DZ twins > 0.02. Finally, 139,072 SNPs with MAF < 0.01 were excluded, leaving 302,627 SNPs for principal component (PC) analysis. The first two PCs of the CATSS-PC samples were projected onto the 1000 Genomes global population reference panel. 236 samples exceeded six standard deviations from the mean values of the European samples in the 1000 Genomes reference, thereby identifying these samples as non-European ancestral outliers. After the removal of the ancestral outliers, a second round of the QC procedure described above was performed, excluding a further seven samples and 23 SNPs. 10789 samples and 302604 SNPs, of which 293590 were successfully aligned to the forward genomic strand and matched to the reference panel, were then used for imputation. The Sanger imputation server was used to impute the post-QC genotype data, using the Haplotype Reference Consortium (HRC v.1.1) (McCarthy et al., 2016) as a reference. EAGLE2 (Loh, Danecek, et al., 2016) was used for pre-phasing and PBWT (Durbin & Barrett, 2014; Rubinacci et al., 2020) was used for imputing. After imputation, 40 million SNPs were available.

For CATSS-GSA, genotypes were generated in six batches during 2018 and 2019 on the Illumina Infinium assay (chip GSAMD-24v1-0_20011747_A1) at the SNP&SEQ Technology Platform at Uppsala University using GenomeStudio 2.0.3. 14 samples were excluded for having sex chromosome abnormalities, genotypic sex different from register information or showing unexpected relatedness patterns, indicating a sample mix-up. Data from genotyped monozygotic twin pairs with genotype missingness < 0.02 were merged into one sample per pair (N = 160). 3898 genotyped samples were processed using Ricopili. SNPs with a missingness higher than 0.05 (N = 11,299) were removed, followed by the exclusion of 8 samples due to either per-sample call rate < 0.98, excessive heterozygosity (FHET outside +/- 0.2), or sex mismatch. 199,312 out of 700,078 markers failed SNP QC due to either per-SNP call rate < 0.98, MAF < 0.01, Hardy-Weinberg disequilibrium ($P < 1x10^{-6}$ in MZ twins and $P < 1x10^{-10}$ in DZ twins), or difference in call rate between MZ twins and DZ twins > 0.02. The least common variant was removed for multiallelic sites encoded as multiple markers with the same position but different alleles. Markers having the same position and alleles were merged into one marker per position (removing 625 SNPs), leaving 500,141 directly genotyped SNPs for analysis. The first two PCs of the CATSS samples were projected onto the 1000 Genomes global population reference panel using plink 1.9. (Chang et al., 2015; Purcell et al., 2007) 233 samples exceeded six standard deviations from the mean values of the European samples in the 1000 Genomes reference, thereby identifying these samples as non-European ancestral outliers. Post-QC genotype files were imputed using the Sanger imputation server, using the Haplotype Reference Consortium (HRC v.1.1) as a reference. EAGLE2 was used for pre-phasing and PBWT was used for imputing. After imputation, 40,359,612 SNPs were available.

PC analysis without an external population reference was performed to generate ancestry covariates for association analysis. The first 20 PCs based on common (MAF => 0.05) genotyped markers in linkage equilibrium (LD; pairwise $R^2$ <= 0.1), excluding known regions of long-range LD, were derived from unrelated individuals, and then projected on the full sample of twins. Relatedness of individuals within (twin pairs) and between different family IDs was estimated using the KING algorithm in PLINK 2.0 (Manichaikul et al., 2010). For DZ twins the expected value was 0.25, for MZ twins 0.5. After QC and imputation, MZ co-twins were imputed from their genotyped siblings. The total sample size including MZ co-twins is 4,824. The number of individuals passing genotype- and phenotype QC is 608.

For STR-YATSS genotypes were generated in six batches during 2018 and 2019 on the Illumina Infinium assay (chip GSAMD-24v1-0_20011747_A1) at the SNP&SEQ Technology Platform at Uppsala University using GenomeStudio 2.0.3. 6 samples were excluded for having sex chromosome abnormalities, genotypic sex different from register information or showing unexpected relatedness patterns, indicating a sample mix up. Data from genotyped monozygotic twin pairs with genotype missingness < 0.02 were merged into one sample per pair (N = 1). 2,719 genotyped samples were processed using Ricopili. SNPs with a missingness higher than 0.05 (N = 8,076) were removed, followed by the exclusion of 2 samples due to either per-sample call rate < 0.98, excessive heterozygosity (FHET outside +/- 0.2), or sex mismatch. 199,113 out of 700,078 markers failed SNP QC due to either per-SNP call rate < 0.98, MAF < 0.01, Hardy-Weinberg disequilibrium ($P < 1x10^{-6}$ in MZ twins and $P < 1x10^{-10}$ in DZ twins), or difference in call rate between DZ twins and MZ twins > 0.02. The least common variant was removed for multiallelic sites encoded as multiple markers with the same position but different alleles. Markers having the same position and alleles were merged into one marker per position (removing 661 SNPs), leaving 500,304 directly genotyped SNPs for analysis. The first two PCs of the CATSS samples were projected onto the 1000 Genomes global population reference panel using plink 1.9 (Chang et al., 2015; Purcell et al., 2007). 63 samples exceeded six standard deviations from the mean values of the European samples in the 1000 Genomes reference, thereby identifying these samples as non-European ancestral outliers. Post-QC genotype files were imputed using the Sanger imputation server, using the Haplotype Reference Consortium (HRC v.1.1) as a reference. EAGLE2 was used for pre-phasing and PBWT was used for imputing. After imputation, 40,359,612 SNPs were available.

PC analysis without an external population reference was performed to generate ancestry covariates for association analysis. The first 20 PCs based on common (MAF => 0.05) genotyped markers in linkage equilibrium (LD; pairwise $R^2$ <= 0.1), excluding known regions of long-range LD, were derived from unrelated individuals and then projected on the full sample of twins. Relatedness of individuals within (twin pairs) and between different family IDs was estimated using the KING algorithm in PLINK 2.0 (Manichaikul et al., 2010). For DZ twins the expected value was 0.25, for MZ twins 0.5. After QC and imputation, MZ co-twins were imputed from their genotyped siblings. The total sample size including MZ co-twins is 3,358. The number of individuals passing genotype- and phenotype QC is 2947.

##### *NTR*

Genotyping was done on multiple platforms over time, namely Perlegen-Affymetrix, Affymetrix 6.0, Affymetrix Axiom, Illumina Human Quad Bead 660, Illumina Omni 1M and Illumina GSA. On each platform genotyping was performed following manufacturers protocols, using the then appropriate calling software. The SNPs of the Perlegen-Affymetrix, Illumina Human Quad Bead 660 and Illumina Omni 1M arrays, which were originally typed on older genome builds, were lifted over to Build 37 HG19 based on RSid locations of the DBSNP 142 marker map. For each genotype platform, samples were removed if DNA sex did not match the expected phenotype, if the Plink heterozygosity F statistic was < -0.10 or > 0.10, or if the genotyping call rate was < 0.90. SNPs were removed if the minor allele frequency (MAF) < 0.01, if the Hardy-Weinberg Equilibrium (HWE) p-value < $1×10^{-5}$, if call rate < 0.95, or if the N Mendel errors > 20 (Purcell et al., 2007). The absolute value of 20 is used here to remove only the worst offending SNPs (N=2902) in platforms that have familial data present (later this is re-filtered more stringently). In addition, palindromic AT/GC SNPs with a MAF range between 0.4 and 0.5 were removed to avoid possible strand alignment issues. For each platform, the data was then position - and strand aligned with the GONL reference set V4. SNPs that had a difference in allele frequency > 0.10 or had mismatching alleles with this reference panel were removed in this step.

The data of the 6 platforms was merged into a single dataset keeping all QCed SNPs of each platform (N = 1,781,526). For each individual only one platform was chosen in the following order: Axiom (3,144) > Affy6 (8,640) > 1M (238) > 660 (1,439) > GSA (5,938) > Affy-Perl (1,238). Based on the ~10.6k SNPs that all platforms have in common, DNA IBD was estimated for all individual pairs using the PLINK and KING programs (Manichaikul et al., 2010; Purcell et al., 2007). These estimates were then compared to the expected familial relations, and samples were removed if these failed to fit. A similar approach was used for DNA zygosity mismatches. Duplicate monozygotic twins N = 3,032, triplets N = 7 as well as NTR samples present in the GONL data N = 364 (plus their MZ-twins) were removed from the data prior to imputation. The data were then cross-platform phased and imputed using MACH-ADMIX to predict the missing SNP genotypes in each platform as compared to the other platforms, based on the complete GONL reference panel haplotypes for the SNPs that were present in at least one platform (the ~1.78m) (Boomsma et al., 2014; Deelen et al., 2014; Fedko et al., 2015; Liu et al., 2013). Post imputation, the 2nd (and 3rd) MZ, plus the GONL samples were re-added duplicating the data from the 1st imputed MZ twin, and the complete SNP data from the GONL reference panel.

After this imputation, SNP QC was redone, now using the full merged dataset with all missing genotypes imputed. SNPs were removed if the HWE p-value was < $1×10^{-5}$, Mendel error rate was more than mean + 3SD, the $R^2$ imputation quality metric was < 0.90 if p-value for association with a single platform vs. all others was < $1×10^{-5}$. No MAF filter was re-applied (min = 0.0025). This left a cleaned merged dataset of 21,001 NTR individuals with 3,032 MZ pairs, 7 MZ trios, and 1,314,639 SNP markers ($N_{chrX}$=20,792). This cross-platform imputed set described is what we consider to be our 'genotyped' dataset in the next two steps, which are the detection of ethnic outliers and the imputation to the 1000 genomes Phase 3v5 and the Human Reference Consortium (HRC) panels (McCarthy et al., 2016; The 1000 Genomes Project Consortium, 2015).

Ancestry outliers (non-Dutch ancestry) were defined based on Principal Components Analysis (PCA) by projecting 10 PCs from 1000G reference set populations on the NTR cross-platform imputed data using the SMARTPCA program as described earlier (Abdellaoui et al., 2013; Price et al., 2006). Individuals with PC values located outside of the range of European and/or British populations were defined as outliers (N=1823). Upon exclusion of outliers, 10 PCs were recomputed for NTR cross-platform imputed data to capture the variation within the Netherlands.

Genotype imputation to the HRC 1.1 (~40m SNPs) and 1000G Phase 3 version 5 (~49m SNPs) reference panels was done on the Michigan Imputation server on the cross-platform imputed data (Das et al., 2016). For each reference panel the data were aligned using the PERL based "HRC or 1000G Imputation preparation and checking" tool v4.2.5 (https://www.well.ox.ac.uk/~wrayner/tools/) .The remaining SNPs (1,302,481: 1000G, 1,307,940: HRC) were then phased with EAGLE for the autosomes, and SHAPEIT for chromosome X and then imputed using Minimach 3 following the standard imputation procedures of the server (Delaneau et al., 2011; Loh, Palamara, et al., 2016).

The cross-chip imputed data was filtered for SNPs having a MAF < 0.01. Then Genetic Relationship Matrices (GRM) were computed for each individual chromosome (1 to 22) using the GCTA software (Yang et al., 2011). Subsequently, these 22 matrices were merged into single autosomal matrix. Leave One Chromosome Out (LOCO) GRMs in order to control for genetic background when running genome wide associations, were calculated likewise by merging 22 subsets of 21 chromosomes respectively. Finally, a specific family based GRM was made by setting all individual pairs sharing less than 0.05 of their genome to 0 in the matrix and leaving the other pairs as calculated (Zaitlen et al., 2013). No GRMs were calculated for the imputed data (100G and HRC) as these are sufficient to control for confounding in GWAS and to calculate the heritability for various traits.

Before association, final filters were applied to the final set of available samples: Ancestry outliers were defined based on Principal Components Analysis (PCA) by projecting 10 PCs from 1000G reference Phase 3v5. We finally filtered on population based and sample MAF filtered at 0.03. Allele-frequency differences between 1000G reference and sample over 0.20 were removed (10,260 SNPs).

##### *SfS*

For detailed methods regarding sample collection, DNA extraction, quantification, and genotyping, as well as data QC refer to (Burton et al., 2021). In brief, saliva samples were collected using the Oragene OG-500 saliva kits (DNA Genotek, Ottawa, Canada). DNA was extracted using standard methods and quantified using real-time polymerase chain reaction (rt-PCR). DNA extracts were genotyped on either the Illumina HumanCoreExome or HumanOmni1 beadchip arrays (Illumina, San Diego, CA, USA). Standard genetic data quality control was conducted in GenomeStudio® and PLINK (Purcell, 2007). Imputation was performed separately for all batches and Illumina platforms using Beagle v4.1 (Browning, 2017) and the phase 3 version 5 data of the 1000 Genomes project as a reference. Subsequent analyses were performed using hard called genotypes with imputation quality (INFO) score >0.8. All non-European ancestry participants were excluded based on principal components analysis (PCA), and only one participant was included from each family (i.e. the first enrolled sibling). For additional exclusion criteria, refer to (Burton et al., 2021).

To generate ancestry related covariates, PCA was performed without an external population reference using a set of SNPs pruned based on linkage disequilibrium (LD; pairwise $r^2$<0.1) and excluding long-range LD regions. To further account for cryptic relatedness in the SfS sample, principal components (PCs) were computed first in unrelated individuals and then projected onto the full SfS cohort to yield projected PCs. For subsequent GWAS and PRS analyses, PCs 1-3 and projected PCs 1-3 were included as covariates.

##### *TwinsUK*

Genotyping of the TwinsUK dataset was done using the Illumina arrays HumanHap300, HumanHap610Q, 1M-Duo and 1.2MDuo 1M. The normalized intensity data for each of the three arrays was pooled separately, with 1M-Duo and 1.2MDuo 1M pooled together. The Illuminus calling algorithm was used to assign genotypes in the pooled data for each dataset. No call was assigned for genotypes with posterior probabilities below a threshold of 0.95. Validation of pooling was achieved via a visual inspection of 100 random, shared SNPs for overt batch effects. Intensity cluster plots of significant SNPs were visually inspected for over-dispersion, biased no-calling, and/or erroneous genotype assignment. SNPs exhibiting any of these characteristics were discarded.

Quality control steps were implemented using PLINK 1.9. Similar exclusion criteria were applied with respect to (i) HumanHap300 and (ii) HumanHap610Q, 1M-Duo, 1.2MDuo 1M genotyped samples). The following sample-wise exclusion criteria were applied: sample call rate < 98%; heterozygosity across all SNPs ≥ 2 SD from the sample mean; evidence of non-European ancestry (this was assessed by PCA comparison with 1000 Genomes by projecting the first two PCs for UKTwins samples onto the 1000 genomes reference populations, using the EIGENSOFT package); observed pairwise IBD probabilities suggestive of sample identity errors. Misclassified monozygotic and dizygotic twins were corrected based on the derived IBD probabilities, calculated using the KING algorithm in PLINK. Ancestry covariates (20 PCs based on MAF => 0.05 and pairwise $R^2$ <= 0.1 after exclusion of long-range LD regions), were derived for the association analysis for unrelated individuals, by re-calculating PCs after having excluded the PC outliers. Pairwise relatedness of individuals was estimated using the KING algorithm in PLINK 2.0 (Manichaikul et al, 2010), using expected values of 0.25 for DZ twins and 0.5, for MZ twins. The following SNP-wise exclusion criteria were applied: Hardy-Weinberg p-value < $10^{-6}$, (assessed using unrelated samples); MAF < 1%, in unrelated samples (iii) SNP call rate < 97% (SNPs with MAF ≥ 5%) or < 99% (for 1% ≤ MAF < 5%). Genotypic data resulting from quality control arms (i) and (ii) were combined and the map was converted from Build36 to Build37.

The HRC/1KG Imputation Preparation and Checking Tool (http://www.well.ox.ac.uk/~wrayner/tools/HRC-1000G-check-bim.v4.2.5.zip) (developed by Will Rayner) was used to check input data for accuracy relative to expected 1000G inputs prior to imputation. This process identified errors in the original data, including incorrect REF/ALT designations, incorrect strand designations, extreme deviations from expected allele frequencies, and palindromic (A/T and G/C) SNPs with allele frequency near 0.5 that are often the source of imputation errors. The problematic variants identified were fixed or removed.

The cleaned/updated binary files (one for each chromosome) generated by this tool were used for phasing, using EAGLE for the autosomes and SHAPEIT for chromosome X. They were then imputed using Minimach 3 following standard procedures (Delaneau et al, 2011, Loh et al, 2016). The 1,000 Genomes Phase 3 integrated variant set (NCBI build 37/hg19 coordinates) served as the reference panel. Hard called genotypes with imputation quality (INFO) score > 0.8 were retained for the downstream analyses.

### 2. Results

#### 2.1. Genome-wide association results

**Supplementary Figure S11**: A) The QQ-plot displays quantiles of the -log(10) p-values, resulting from the inverse weighted GWAS meta-analysis, plotted against the quantiles expected under the null hypothesis. 95% confidence interval is indicated by the grey shading. The genomic inflation factor Lambda is the observed median Chi² test statistic under the null hypothesis. Lambda1000 indicates the lambda if the sample contains 1000 individuals. Number of SNPs (N(pvals)) are the number of SNPS included in the meta-analysis. B) The QQ-plot from the gene-based analysis, displaying quantiles of the -log(10) p-values plotted against the quantiles expected under the null hypothesis.

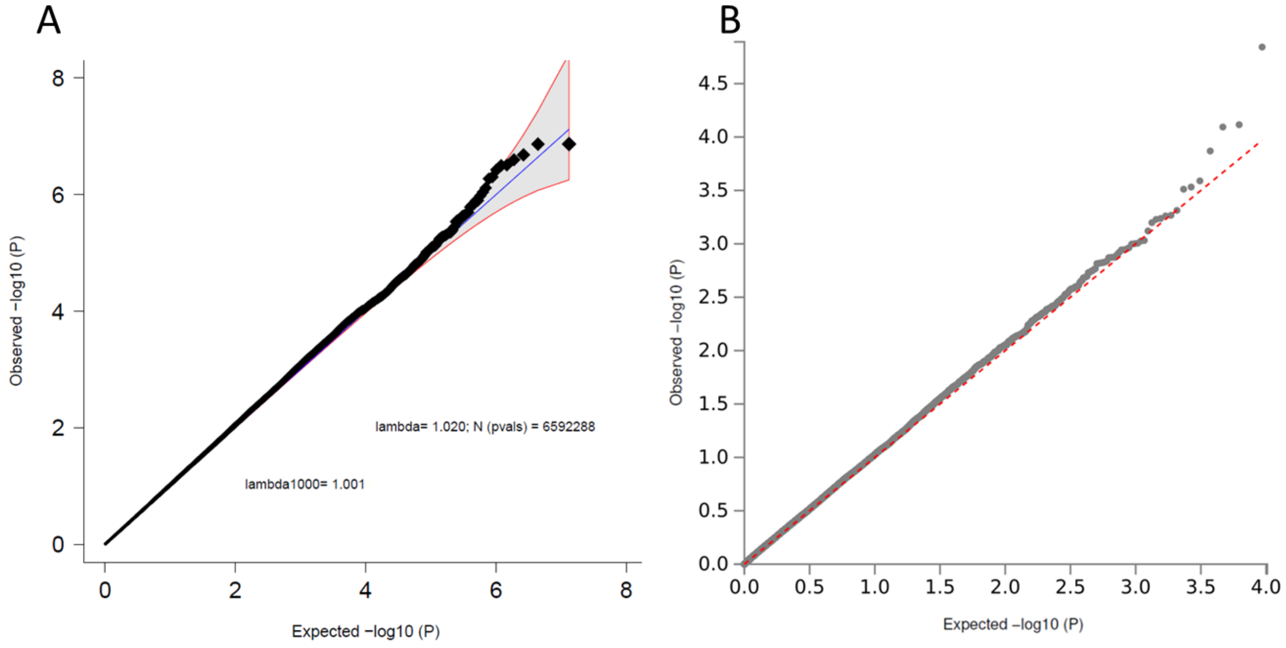

**Supplementary Figure S12**: Forestplot (A) and regional association plot (B) of SNP rs1173221479. A) The forestplot shows the effect estimate with 95%-confidence interval for each cohort contributing to the meta-analysis and for the inverse variance weighted meta-analysis (bottom line). The table lists INFO (imputation quality score), p-value (p-value of the SNP effect), f_ca(n) (frequency cases), f_co(n) (frequency controls) (as HS is a quantitative phenotype, f_ca(n) and f_co(n) are both the same and the overall frequency), ln(OR) (Beta estimate of the effect of the SNP), and STDerr (standard error of ln(OR)) for each of the contributing cohorts and for the meta-analysis. At the top, '+' indicates a positive direction of effect, '-' a negative direction of effect, while '?' indicates that the SNP was not contained in the respective cohort. B) The –log10(p-value) of SNPs in the HS meta-analysis GWAS is shown on the left y-axis. The recombination rates expressed in centimorgans (cM) per Mb (Megabase) (light blue line) are shown on the right y-axis. Position in Mb is on the x axis. Only the SNPs with an association p-value less than 0.1 were plotted. The most associated SNP is shown as a purple diamond.

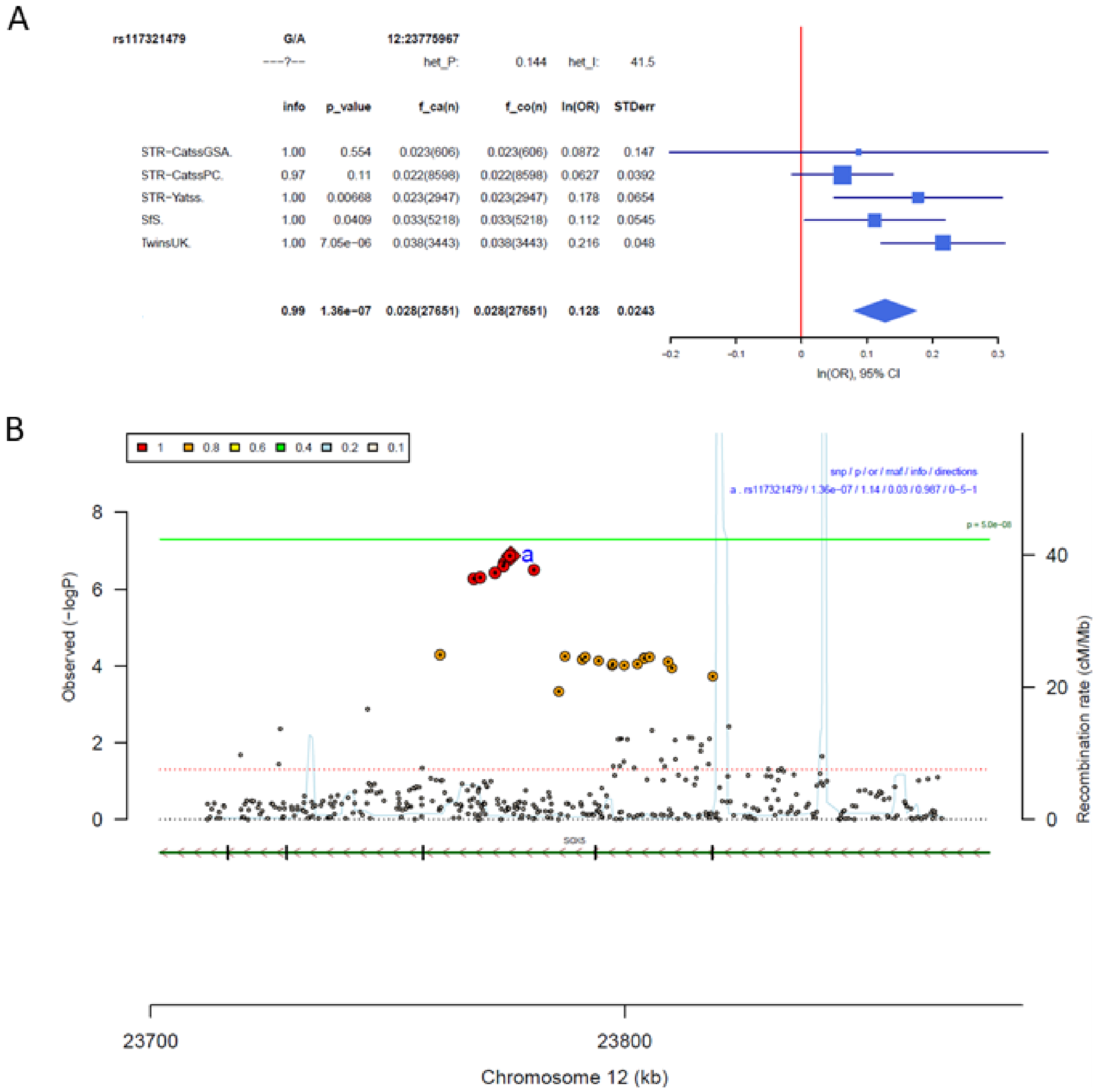

**Supplementary Figure S13:** Forestplot (A) and regional association plot (B) of SNP rs78426839. A) The forestplot shows the effect estimate with 95%-confidence interval for each cohort contributing to the meta-analysis and for the inverse variance weighted meta-analysis (bottom line). The table lists INFO (imputation quality score), p-value (p-value of the SNP effect), f_ca(n) (frequency cases), f_co(n) (frequency controls) (as HS is a quantitative phenotype, f_ca(n) and f_co(n) are both the same and the overall frequency), ln(OR) (Beta estimate of the effect of the SNP), and STDerr (standard error of ln(OR)) for each of the contributing cohorts and for the meta-analysis. At the top, '+' indicates a positive direction of effect, '-' a negative direction of effect, while '?' indicates that the SNP was not contained in the respective cohort. B) The –log10(p-value) of SNPs in the HS meta-analysis GWAS is shown on the left y-axis. The recombination rates expressed in centimorgans (cM) per Mb (Megabase) (light blue line) are shown on the right y-axis. Position in Mb is on the x axis. Only the SNPs with an association p-value less than 0.1 were plotted. The most associated SNP is shown as a purple diamond.

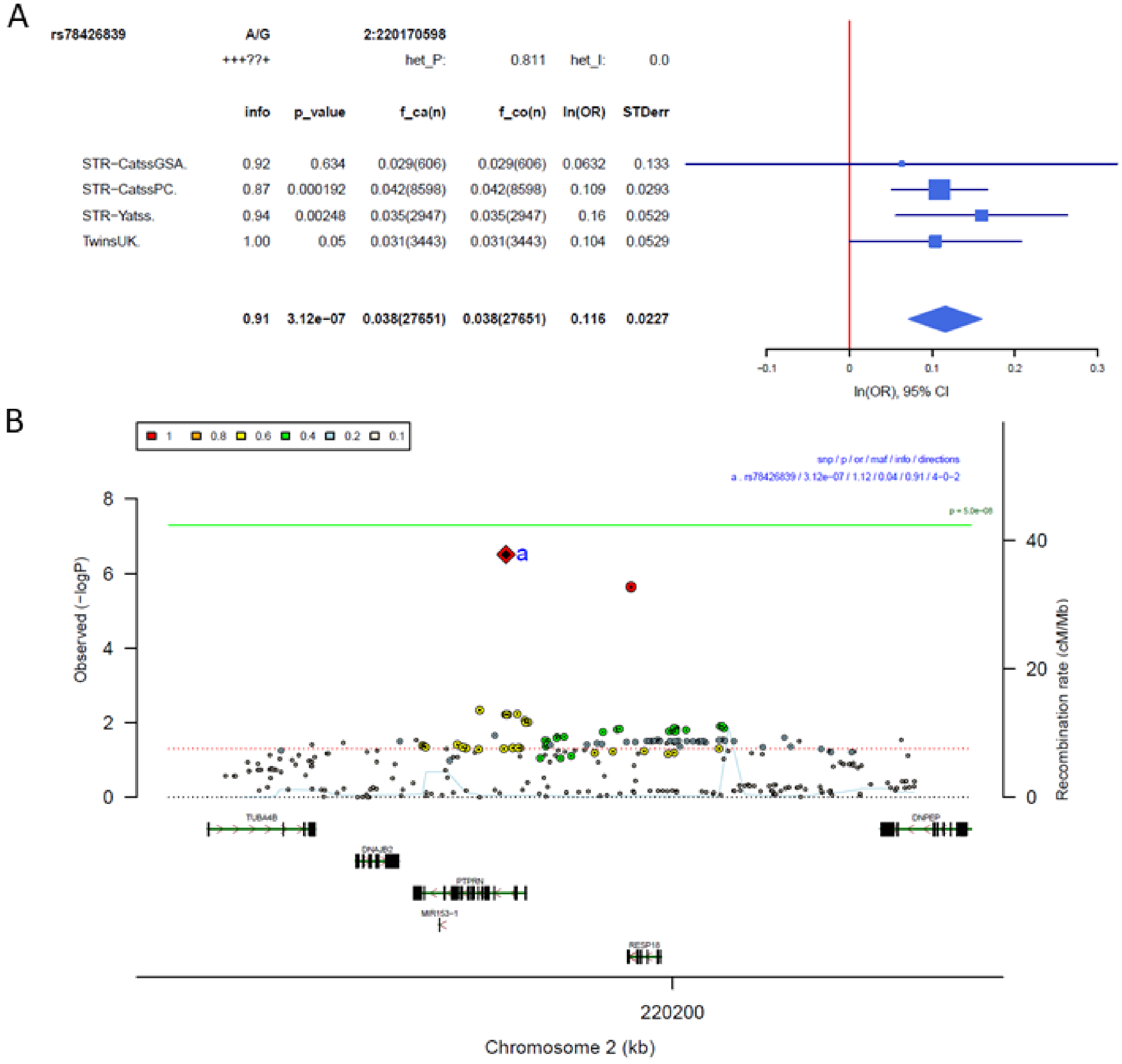

**Supplementary Figure S14:** Forestplot (A) and regional association plot (B) of SNP rs7567224. A) The forestplot shows the effect estimate with 95%-confidence interval for each cohort contributing to the meta-analysis and for the inverse variance weighted meta-analysis (bottom line). The table lists INFO (imputation quality score), p-value (p-value of the SNP effect), f_ca(n) (frequency cases), f_co(n) (frequency controls) (as HS is a quantitative phenotype, f_ca(n) and f_co(n) are both the same and the overall frequency), ln(OR) (Beta estimate of the effect of the SNP), and STDerr (standard error of ln(OR)) for each of the contributing cohorts and for the meta-analysis. At the top, '+' indicates a positive direction of effect, '-' a negative direction of effect, while '?' indicates that the SNP was not contained in the respective cohort. B) The –log10(p-value) of SNPs in the HS meta-analysis GWAS is shown on the left y-axis. The recombination rates expressed in centimorgans (cM) per Mb (Megabase) (light blue line) are shown on the right y-axis. Position in Mb is on the x axis. Only the SNPs with an association p-value less than 0.1 were plotted. The most associated SNP is shown as a purple diamond.

A

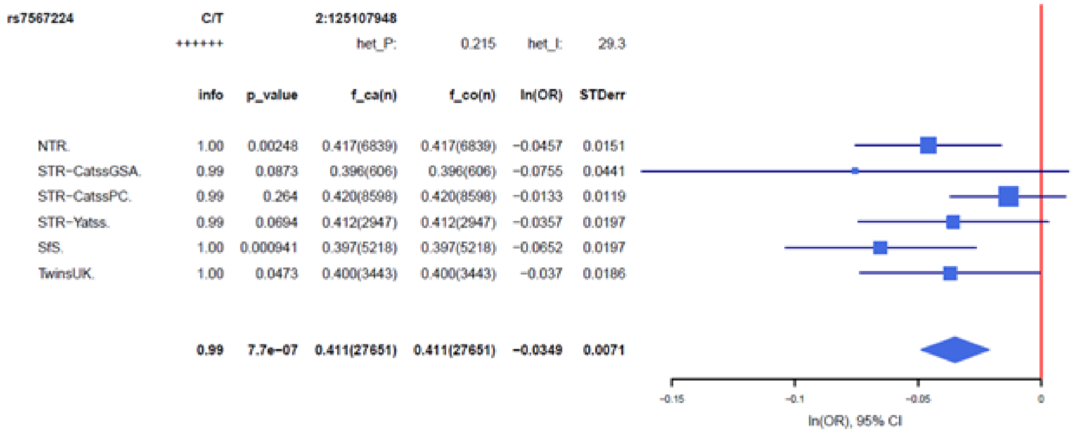


| | info | p_value | f_ca(n) | f_co(n) | ln(OR) | STDerr |
|---|---|---|---|---|---|---|
| NTR. | 1.00 | 0.00248 | 0.417(6839) | 0.417(6839) | −0.0457 | 0.0151 |
| STR−CatssGSA. | 0.99 | 0.0873 | 0.396(606) | 0.396(606) | −0.0755 | 0.0441 |
| STR−CatssPC. | 0.99 | 0.264 | 0.420(8598) | 0.420(8598) | −0.0133 | 0.0119 |
| STR−Yatss. | 0.99 | 0.0694 | 0.412(2947) | 0.412(2947) | −0.0357 | 0.0197 |
| SfS. | 1.00 | 0.000941 | 0.397(5218) | 0.397(5218) | −0.0652 | 0.0197 |
| TwinsUK. | 1.00 | 0.0473 | 0.400(3443) | 0.400(3443) | −0.037 | 0.0186 |
| | **0.99** | **7.7e−07** | **0.411(27651)** | **0.411(27651)** | **−0.0349** | **0.0071** |

B

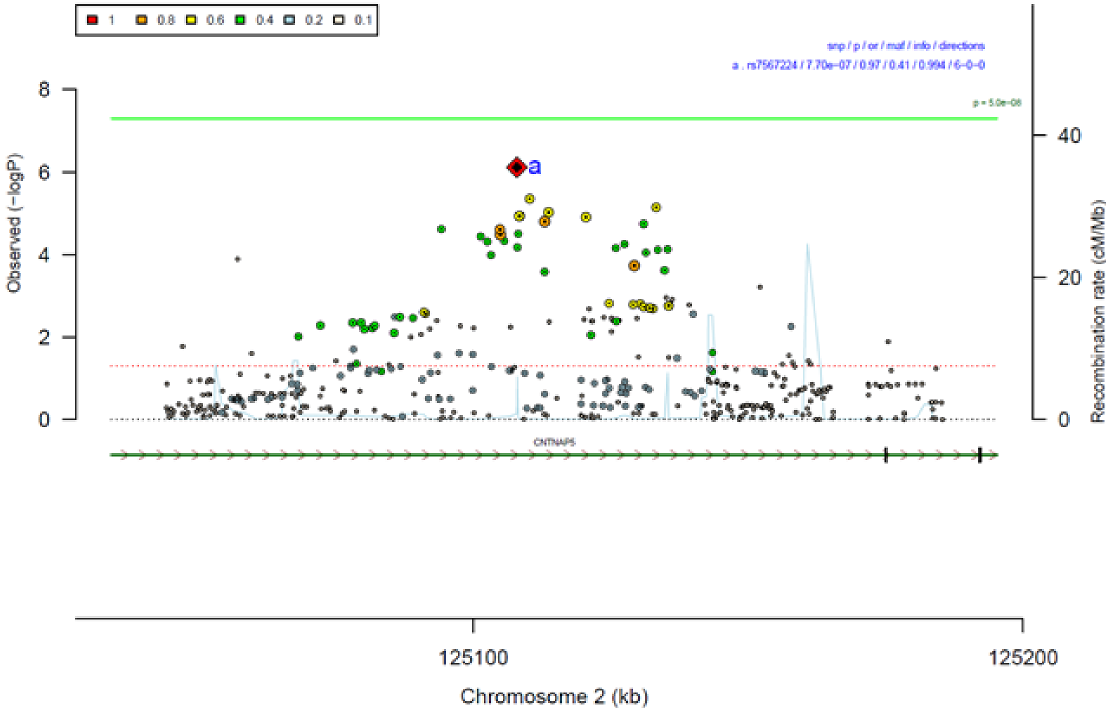

**Supplementary Figure S15:** Forestplot (A) and regional association plot (B) of SNP rs72927972. A) The forestplot shows the effect estimate with 95%-confidence interval for each cohort contributing to the meta-analysis and for the inverse variance weighted meta-analysis (bottom line). The table lists INFO (imputation quality score), p-value (p-value of the SNP effect), f_ca(n) (frequency cases), f_co(n) (frequency controls) (as HS is a quantitative phenotype, f_ca(n) and f_co(n) are both the same and the overall frequency), ln(OR) (Beta estimate of the effect of the SNP), and STDerr (standard error of ln(OR)) for each of the contributing cohorts and for the meta-analysis. At the top, ‘+’ indicates a positive direction of effect, ‘-’ a negative direction of effect, while ‘?’ indicates that the SNP was not contained in the respective cohort. B) The –log10(p-value) of SNPs in the HS meta-analysis GWAS is shown on the left y-axis. The recombination rates expressed in centimorgans (cM) per Mb (Megabase) (light blue line) are shown on the right y-axis. Position in Mb is on the x axis. Only the SNPs with an association p-value less than 0.1 were plotted. The most associated SNP is shown as a purple diamond.

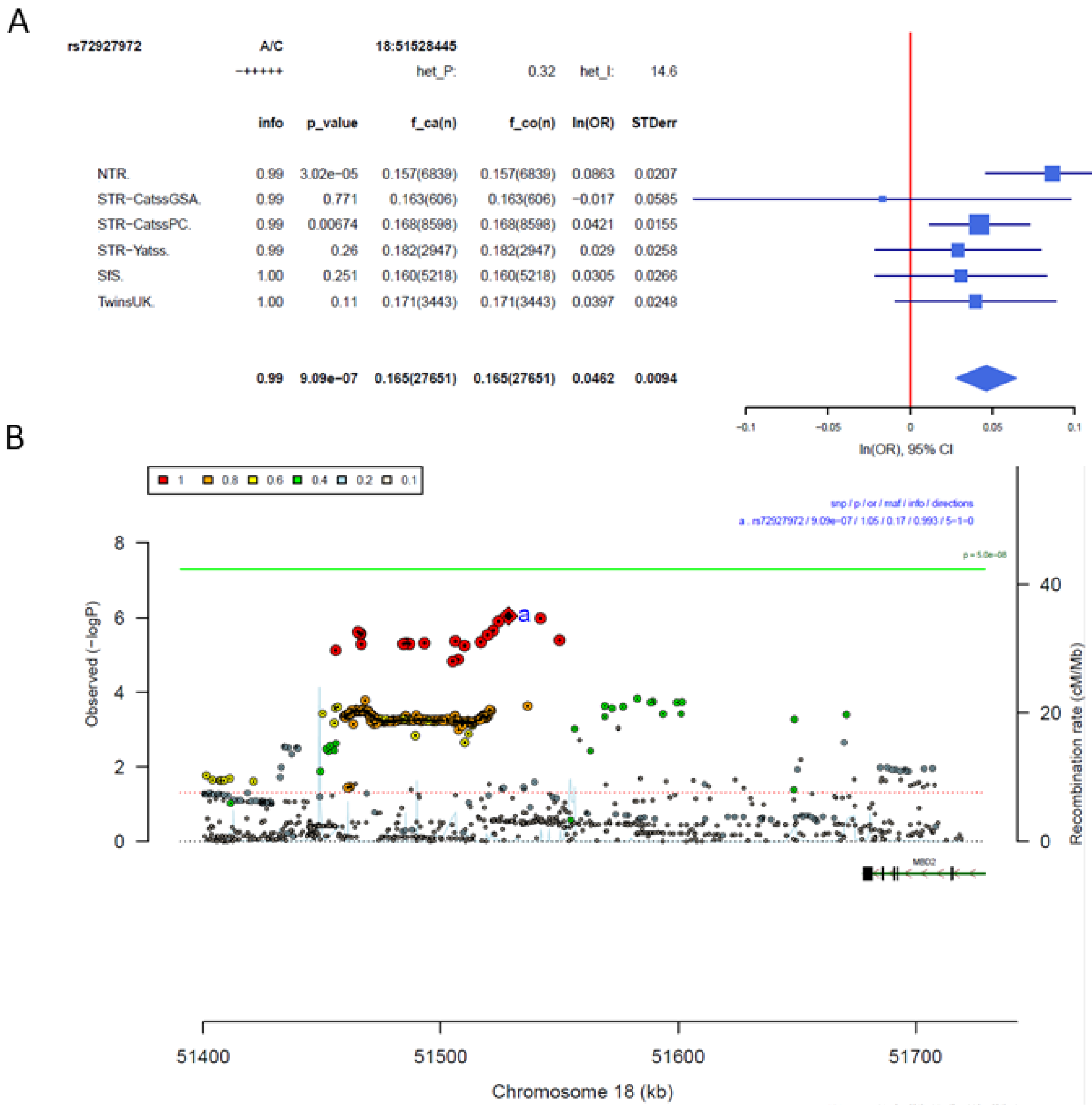

#### 2.2. Compatibility between cohorts

**Supplementary Figure S16:** Manhattanplot and qqplot of heterogeneity test

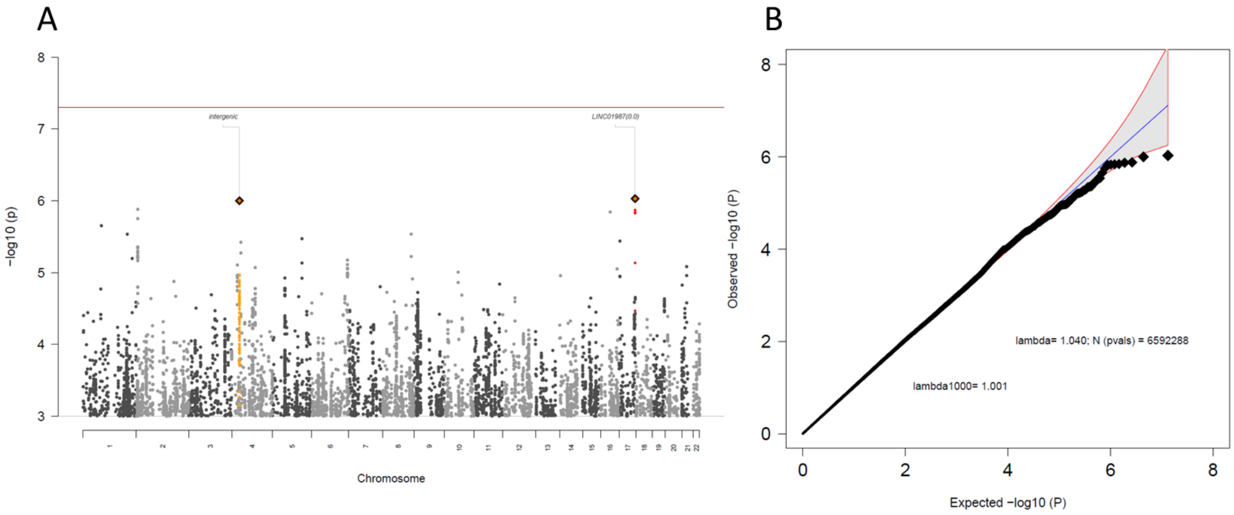

### Supplementary References


Abdellaoui, A., Hottenga, J. J., Knijff, P. De, Nivard, M. G., Xiao, X., Scheet, P., Brooks, A., Ehli, E. A., Hu, Y., Davies, G. E., Hudziak, J. J., Sullivan, P. F., Van Beijsterveldt, T., Willemsen, G., De Geus, E. J., Penninx, B. W. J. H., & Boomsma, D. I. (2013). Population structure, migration, and diversifying selection in the Netherlands. *European Journal of Human Genetics : EJHG*, *21*(11), 1277–1285. https://doi.org/10.1038/EJHG.2013.48

Ameis, S. H., Lerch, J. P., Taylor, M. J., Lee, W., Viviano, J. D., Pipitone, J., Nazeri, A., Croarkin, P. E., Voineskos, A. N., Lai, M. C., Crosbie, J., Brian, J., Soreni, N., Schachar, R., Szatmari, P., Arnold, P. D., & Anagnostou, E. (2016). A diffusion tensor imaging studyin children with ADHD, autism spectrum disorder, OCD, and matched controls: Distinct and non-distinct white matter disruption and dimensional brain-behavior relationships. *American Journal of Psychiatry*, *173*(12), 1213–1222. https://doi.org/10.1176/appi.ajp.2016.15111435

Anckarsäter, H., Lundström, S., Kollberg, L., Kerekes, N., Palm, C., Carlström, E., Långström, N., Magnusson, P. K. E., Halldner, L., Bölte, S., Gillberg, C., Gumpert, C., Råstam, M., & Lichtenstein, P. (2011). The Child and Adolescent Twin Study in Sweden (CATSS). *Twin Research and Human Genetics : The Official Journal of the International Society for Twin Studies*, *14*(6), 495–508. https://doi.org/10.1375/twin.14.6.495

Andrew, T., Hart, D. J., Snieder, H., De Lange, M., Spector, T. D., & Macgregor, A. J. (2001). Are twins and singletons comparable? A study of disease-related and lifestyle characteristics in adult women. *Twin Research : The Official Journal of the International Society for Twin Studies*, *4*(6), 464–477. https://doi.org/10.1375/1369052012803

Boomsma, D. I., Vink, J. M., Beijsterveldt, T. C. E. . M. van, Geus, E. J. C. de, Beem, A. L., Mulder, E. J. C. M., Derks, E. M., Riese, H., Willemsen, G. A. H. M., Bartels, M., Berg, M. van den, Kupper, N. H. M., Polderman, T. J. C., Posthuma, D., Rietveld, M. J. H., Stubbe, J. H., Knol, L. I., Stroet, T., & Baal, G. C. M. van. (2002). Netherlands Twin Register: A Focus on Longitudinal Research. *Twin Research and Human Genetics*, *5*(5), 401–406. https://doi.org/10.1375/TWIN.5.5.401

Boomsma, D. I., Wijmenga, C., Slagboom, E. P., Swertz, M. A., Karssen, L. C., Abdellaoui, A., Ye, K., Guryev, V., Vermaat, M., Van Dijk, F., Francioli, L. C., Hottenga, J. J., Laros, J. F. J., Li, Q., Li, Y., Cao, H., Chen, R., Du, Y., Li, N., … Van Duijn, C. M. (2014). The Genome of the Netherlands: design, and project goals. *European Journal of Human Genetics*, *22*(2), 221–227. https://doi.org/10.1038/EJHG.2013.118

Chang, C. C., Chow, C. C., Tellier, L. C. A. M., Vattikuti, S., Purcell, S. M., & Lee, J. J. (2015). Second-generation PLINK: Rising to the challenge of larger and richer datasets. *GigaScience*, *4*(1), 7. https://doi.org/10.1186/S13742-015-0047-8/2707533

Crosbie, J., Arnold, P., Paterson, A., Swanson, J., Dupuis, A., Li, X., Shan, J., Goodale, T., Tam, C., Strug, L. J., & Schachar, R. J. (2013). Response Inhibition and ADHD Traits: Correlates and Heritability in a Community Sample. *Journal of Abnormal Child Psychology*, *41*(3), 497. https://doi.org/10.1007/S10802-012-9693-9

Das, S., Forer, L., Schönherr, S., Sidore, C., Locke, A. E., Kwong, A., Vrieze, S. I., Chew, E. Y., Levy, S., McGue, M., Schlessinger, D., Stambolian, D., Loh, P. R., Iacono, W. G., Swaroop, A., Scott, L. J., Cucca, F., Kronenberg, F., Boehnke, M., … Fuchsberger, C. (2016). Next-generation genotype imputation service and methods. *Nature Genetics*, *48*(10), 1284–1287. https://doi.org/10.1038/NG.3656

Deelen, P., Menelaou, A., Van Leeuwen, E. M., Kanterakis, A., Van Dijk, F., Medina-Gomez, C., Francioli, L. C., Hottenga, J. J., Karssen, L. C., Estrada, K., Kreiner-Møller, E., Rivadeneira, F., Van Setten, J., Gutierrez-Achury, J., Westra, H. J., Franke, L., Van Enckevort, D., Dijkstra, M., Byelas, H., … Swertz, M. A. (2014). Improved imputation quality of low-frequency and rare variants in European samples using the "Genome of The Netherlands." *European Journal of Human Genetics*, *22*(11), 1321–1326. https://doi.org/10.1038/EJHG.2014.19

Delaneau, O., Marchini, J., & Zagury, J. F. (2011). A linear complexity phasing method for thousands of genomes. *Nature Methods 2011 9:2*, *9*(2), 179–181. https://doi.org/10.1038/nmeth.1785

Durbin, R., & Barrett, J. (2014). Efficient haplotype matching and storage using the positional Burrows–Wheeler transform (PBWT). *Bioinformatics*, *30*(9), 1266–1272. https://doi.org/10.1093/BIOINFORMATICS/BTU014

Fedko, I. O., Hottenga, J. J., Medina-Gomez, C., Pappa, I., van Beijsterveldt, C. E. M., Ehli, E. A., Davies, G. E., Rivadeneira, F., Tiemeier, H., Swertz, M. A., Middeldorp, C. M., Bartels, M., & Boomsma, D. I. (2015). Estimation of Genetic Relationships Between Individuals Across Cohorts and Platforms: Application to Childhood Height. *Behavior Genetics*, *45*(5), 514–528. https://doi.org/10.1007/S10519-015-9725-7

Ivanov, V. Z., Nordsletten, A., Mataix-Cols, D., Serlachius, E., Lichtenstein, P., Lundström, S., Magnusson, P. K. E., Kuja-Halkola, R., & Rück, C. (2017). Heritability of hoarding symptoms across adolescence and young adulthood: A longitudinal twin study. *PloS One*, *12*(6). https://doi.org/10.1371/JOURNAL.PONE.0179541

Lam, M., Awasthi, S., Watson, H. J., Goldstein, J., Panagiotaropoulou, G., Trubetskoy, V., Karlsson, R., Frei, O., Fan, C. C., De Witte, W., Mota, N. R., Mullins, N., Brügger, K., Hong Lee, S., Wray, N. R., Skarabis, N., Huang, H., Neale, B., Daly, M. J., … Ripke, S. (2020). RICOPILI: Rapid Imputation for COnsortias PIpeLIne. *Bioinformatics*, *36*(3), 930–933. https://doi.org/10.1093/bioinformatics/btz633

Liu, E. Y., Li, M., Wang, W., & Li, Y. (2013). MaCH-Admix: Genotype Imputation for Admixed Populations. *Genetic Epidemiology*, *37*(1), 25–37. https://doi.org/10.1002/gepi.21690

Loh, P. R., Danecek, P., Palamara, P. F., Fuchsberger, C., Reshef, Y. A., Finucane, H. K., Schoenherr, S., Forer, L., McCarthy, S., Abecasis, G. R., Durbin, R., & Price, A. L. (2016). Reference-based phasing using the Haplotype Reference Consortium panel. *Nature Genetics 2016 48:11*, *48*(11), 1443–1448. https://doi.org/10.1038/ng.3679

Loh, P. R., Palamara, P. F., & Price, A. L. (2016). Fast and accurate long-range phasing in a UK Biobank cohort. *Nature Genetics 2016 48:7*, *48*(7), 811–816. https://doi.org/10.1038/ng.3571

Manichaikul, A., Mychaleckyj, J. C., Rich, S. S., Daly, K., Sale, M., & Chen, W. M. (2010). Robust relationship inference in genome-wide association studies. *Bioinformatics*, *26*(22), 2867–2873. https://doi.org/10.1093/BIOINFORMATICS/BTQ559

McCarthy, S., Das, S., Kretzschmar, W., Delaneau, O., Wood, A. R., Teumer, A., Kang, H. M., Fuchsberger, C., Danecek, P., Sharp, K., Luo, Y., Sidore, C., Kwong, A., Timpson, N., Koskinen, S., Vrieze, S., Scott, L. J., Zhang, H., Mahajan, A., … Marchini, J. (2016). A reference panel of 64,976 haplotypes for genotype imputation. *Nature Genetics 2016 48:10*, *48*(10), 1279–1283. https://doi.org/10.1038/ng.3643

Price, A. L., Patterson, N. J., Plenge, R. M., Weinblatt, M. E., Shadick, N. A., & Reich, D. (2006). Principal components analysis corrects for stratification in genome-wide association studies. *Nature Genetics*, *38*(8), 904–909. https://doi.org/10.1038/NG1847

Purcell, S., Neale, B., Todd-Brown, K., Thomas, L., Ferreira, M. A. R., Bender, D., Maller, J., Sklar, P., De Bakker, P. I. W., Daly, M. J., & Sham, P. C. (2007). PLINK: a tool set for whole-genome association and population-based linkage analyses. *American Journal of Human Genetics*, *81*(3), 559–575. https://doi.org/10.1086/519795

Rubinacci, S., Delaneau, O., & Marchini, J. (2020). Genotype imputation using the Positional Burrows Wheeler Transform. *PLOS Genetics*, *16*(11), e1009049. https://doi.org/10.1371/JOURNAL.PGEN.1009049

Sarna, S., Kaprio, J., Sistonen, P., & Koskenvuo, M. (1978). Diagnosis of twin zygosity by mailed questionnaire. *Human Heredity*, *28*(4), 241–254. https://doi.org/10.1159/000152964

Spector, T. D., & Williams, F. M. K. (2006). The UK Adult Twin Registry (TwinsUK). *Twin Research and Human Genetics : The Official Journal of the International Society for Twin Studies*, *9*(6), 899–906. https://doi.org/10.1375/183242706779462462

The 1000 Genomes Project Consortium. (2015). A global reference for human genetic variation. *Nature*, *526*(7571), 68–74. https://doi.org/10.1038/nature15393

Yang, J., Lee, S. H., Goddard, M. E., & Visscher, P. M. (2011). GCTA: A tool for genome-wide complex trait analysis. *Am J Hum Genet*, *88*(1), 76–82. https://doi.org/10.1016/j.ajhg.2010.11.011

Zagai, U., Lichtenstein, P., Pedersen, N. L., & Magnusson, P. K. E. (2019). The Swedish Twin Registry: Content and Management as a Research Infrastructure. *Twin Research and Human Genetics : The Official Journal of the International Society for Twin Studies*, *22*(6), 672–680. https://doi.org/10.1017/THG.2019.99

Zaitlen, N., Kraft, P., Patterson, N., Pasaniuc, B., Bhatia, G., Pollack, S., & Price, A. L. (2013). Using Extended Genealogy to Estimate Components of Heritability for 23 Quantitative and Dichotomous Traits. *PLOS Genetics*, *9*(5), e1003520. https://doi.org/10.1371/JOURNAL.PGEN.1003520
